# Expanded T cell clones with lymphoma driver somatic mutations in refractory celiac disease

**DOI:** 10.1101/2024.03.17.24304320

**Authors:** Mandeep Singh, Raymond H. Y. Louie, Jerome Samir, Matthew A. Field, Claire Milthorpe, Thiruni Aldiriki, Joseph Mackie, Ellise Roper, Megan Faulks, Katherine J. L. Jackson, Andrew Calcino, Melinda Y. Hardy, Piers Blombery, Timothy G. Amos, Ira W. Deveson, Scott A. Read, Dmitri Shek, Antoine Guerin, Cindy S Ma, Stuart G. Tangye, Antonio Di Sabatino, Marco V. Lenti, Alessandra Pasini, Rachele Ciccocioppo, Golo Ahlenstiel, Dan Suan, Jason A. Tye-Din, Christopher C. Goodnow, Fabio Luciani

## Abstract

Intestinal inflammation continues in a subset of celiac disease (CD) patients despite a gluten-free diet. Here, by applying multiomic single cell analysis to duodenal biopsies, we find low-grade malignancies with lymphoma driver mutations in refractory CD type 2 (RCD2) patients comprise surface CD3 negative (sCD3-) lymphocytes stalled at an innate lymphoid cell (ILC) - progenitor T cell stage undergoing extensive *TCR* recombination. In people with refractory CD type 1 (RCD1), who currently lack explanation, we discover sCD3+ T cells with lymphoma driver mutations forming large clones displaying inflammatory and cytotoxic molecular profiles in 6 of 10 individuals, and a single small clone in 1 of 4 active recently diagnosed CD cases. Accumulation of driver-mutated T cells and their sCD3-progenitors may explain chronic, non-responsive autoimmunity.

**One-Sentence Summary:** Treatment refractory autoimmunity in celiac disease may be explained by dysregulated T cells and progenitors that have acquired lymphoma-driver mutations.

## Main text

Autoimmune diseases that continue unabated pose a clinical and conceptual challenge. Celiac disease (CD), which affects 0.5%-1.5% of people globally (*1*), is unique among autoimmune disorders because it has a known environmental driver, dietary gluten from wheat, rye and barley (*2, 3*). Deamidation of gluten peptides by tissue transglutaminase promotes binding to HLA-DQ2 and HLA-DQ8 gene products for recognition by T cell receptors (TCRs) on proinflammatory CD4+ T cells in the small intestine (*4*). The immune response is amplified by capture and presentation of the deamidated peptides by intestinal plasma cells making autoantibodies to tissue transglutaminase 2 (TG2) (*2*) and by clonal expansion of CD8 T cells adopting an NK-like cytolytic phenotype (*5–8*). Treatment with a strict, lifelong gluten-free diet generally induces symptom resolution, normalisation of CD serology and healing of the enteropathy but up to 30% of patients experience non-responsive CD (*9, 10*).

A clinically severe form of non-responsive CD, refractory CD (RCD), affects 0.3-4% of CD patients (*9*) and its definition encompasses persistent or recurrent enteropathy in conjunction with malabsorption characterised by diarrhea, abdominal pain, weight loss and anemia, despite more than 12 months on a strict gluten-free diet, with other causes of villous atrophy and malabsorption excluded (*11*). RCD is divided into a common type 1 (RCD1) variant, where the population of duodenal intraepithelial lymphocytes (IEL) appears normal, and a rarer type 2 (RCD2) variant.

RCD2 is diagnosed and treated as low-grade malignancy. The intestinal epithelium is infiltrated with >20% of aberrant lymphocytes expressing intracellular CD3 but lacking surface expression of T cell markers sCD3, CD4 and CD8 (*12–14*). These cells typically carry a monoclonal rearrangement of the TCRγ gene but incomplete or out-of-frame TCRδ and TCRβ gene rearrangements (*15–17*). While they appear unable to differentiate into sCD3+ T cells they resemble a rare subset of thymus-derived, intestinal ILCs that can differentiate into NK cells or sCD3+ T cells in response to NOTCH and IL-15 (*15, 18, 19*). The presence of these aberrant sCD3-lymphocytes defines RCD2, explains the failure to respond to a gluten-free diet, and provides diagnostic markers to inform a different treatment approach. While the aberrant sCD3-lymphocytes in RCD2 are mostly non-cycling, they have multiple somatic mutations considered to be drivers of lymphoma/leukemia (*20, 21*), and evolve into enteropathy-associated T-cell lymphoma (EATL) (*22*), an aggressive intestinal lymphoma with poor prognosis, in 60-80% of RCD2 individuals within 5 years (*23–25*). RCD2 is currently treated with chemotherapy or autologous bone marrow transplant.

In stark contrast to RCD2, the cause of RCD1 is unknown and because RCD1 lacks unique pathological features, the diagnosis is one of exclusion. Differentiating RCD1 from active CD caused by ongoing gluten ingestion in the absence of a biomarker is challenging as both can manifest similarly and identifying trace or inadvertent gluten exposure is difficult. Once RCD1 has been diagnosed, there are limited treatment options other than broad immunosuppression. Thus, there is an urgent unmet need for a mechanistic explanation of RCD1 to inform positive diagnostic indicators and reveal novel treatment targets.

### Intestinal T cells with driver mutations in people with refractory celiac disease

To overcome the limitations of pooled cell DNA sequencing employed by previous studies that identified somatic mutations in RCD2 but not RCD1 nor in uncomplicated CD (*20, 21*), we brought together recent advances in single-cell DNA, RNA and antibody-tag sequencing technology to analyse at much greater resolution the clonal architecture of infiltrating intestinal cells in RCD1, RCD2 and recently diagnosed active CD.

We tested for lymphoma driver mutations in subsets of lymphocytes isolated from duodenal biopsies of 10 individuals with RCD1 compared to 4 individuals with active, newly-diagnosed CD (ANCD) on a gluten-containing diet. As positive and negative controls, we tested biopsies from 2 individuals with RCD2 and 7 people who proved not to have active CD (table S1). The two RCD2 individuals had aberrant sCD3-lymphocytes based on immunohistochemistry and flow cytometry showing >20% CD45^+^ CD103^+^ sCD3^−^ intracellular CD3^+^ (cytCD3^+^) CD4^−^ CD8^−^ intraepithelial populations (data S1, table S1, supplementary text). The 10 RCD1 individuals did not display abnormal frequencies of aberrant sCD3-CD103+ lymphocytes. All RCD1 and ANCD individuals had Marsh scores of 3, indicative of increased lymphocytic infiltration of the epithelium and substantial villous atrophy, while the negative controls had a Marsh score of 0, consistent with normal mucosa (table S1).

A mean of 2085 cells in each biopsy were simultaneously analysed for somatic mutations by single-cell DNA sequencing with targeted amplicons (scDNA-Seq, MissionBio Tapestri) and for 83 cell surface proteins detected with barcoded antibodies (Fig. 1A). The main immune and epithelial cell types in the gut were resolved by combining unsupervised clustering and supervised annotation using canonical markers (Fig. 1B), while removing dead cells, doublets and ambiguous cell types (fig. S1A to E, data S2, see Methods).

**Fig. 1.**
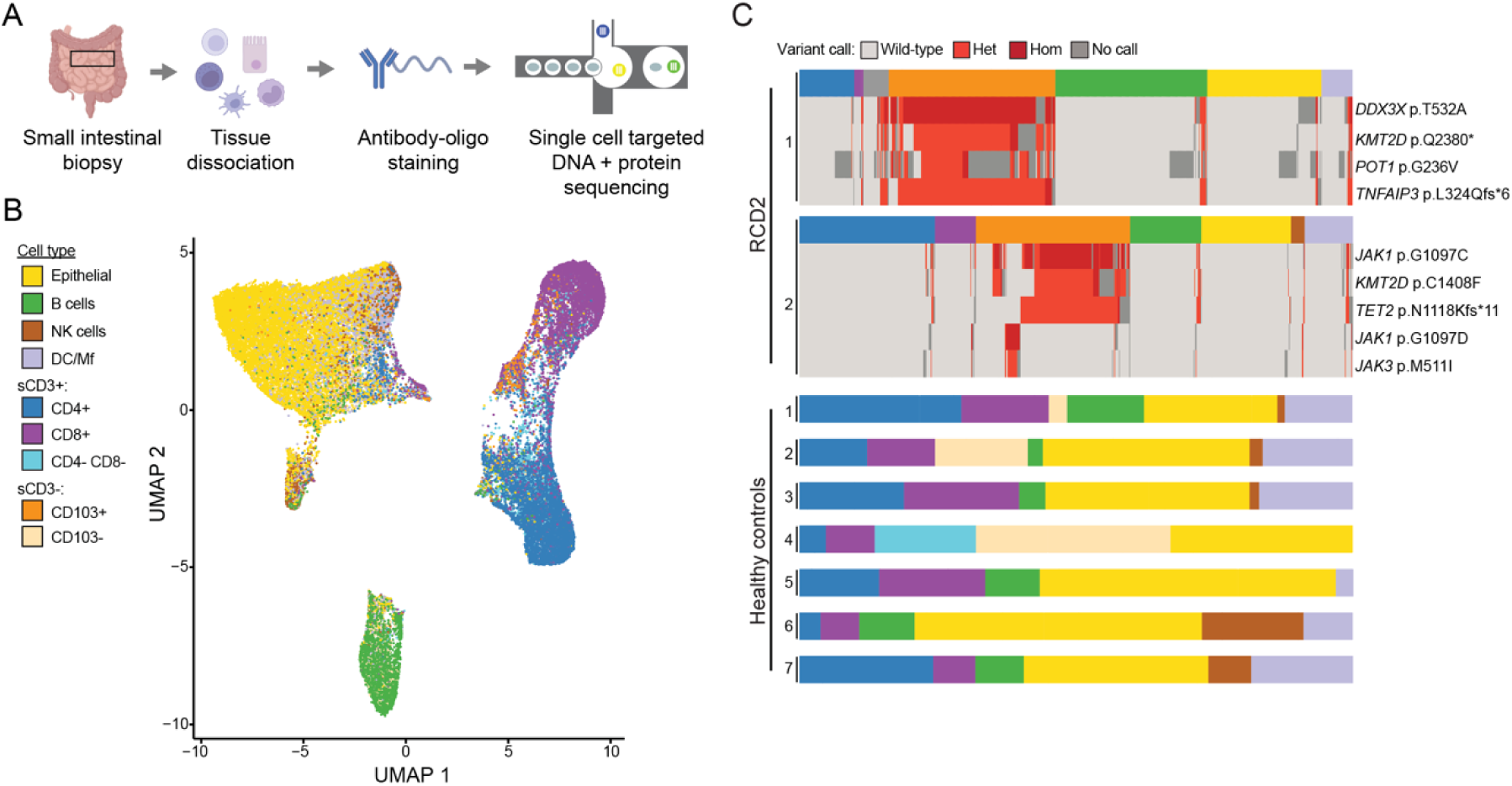
Single cell somatic DNA mutation detection in sCD3-CD103+ cells in RCD2 individuals. **(A)** Strategy to simultaneously analyse cell surface proteins and DNA mutations from thousands of individuals cells obtained from duodenal biopsies (scDNA-Seq). (**B**) Uniform manifold approximation and projection (UMAP) of cells obtained from duodenal biopsies analysed by cell surface proteins of 23 individuals (RCD2, n=2; RCD1, n=10; ANCD, n=4; healthy controls, n=7) integrated together (n= 47,946 cells), and annotated by cell type (15,032 epithelial cells, 5,252 B cells, 7,645 sCD3+ CD8+ cells, 6,912 sCD3+ CD4+ cells, 3,263 sCD3+ CD4-CD8-cells, 798 sCD3-CD103+ cells, 963 sCD3-CD103-cells, 2,174 NK cells, and 5,589 DC and Macrophages (DC/Mf)). (**C**) Boxplots of 1,841 cells from 2 RCD2 subjects and 16,809 cells from 7 healthy control subjects, with coloured segments denoting cells assigned to the indicated cell types (per key in B), and the indicated DNA somatic mutation calls for each cell in rows below for subjects where driver mutations were identified. Mutations called by scDNA-Seq as either wild-type, heterozygous (Het), homozygous (Hom), or no call are denoted by different colours as per the key. The subject identifier is depicted at the left of each boxplot.

Utilising this approach, we profiled 47,946 cells across all individuals including 18,138 sCD3+ T cells (7,645 CD8+ and 6,912 CD4+); 5,252 B cells; 15,032 epithelial cells; 5,589 dendritic cells / macrophages; 2,174 NK cells; and 1,761 sCD3-lymphocytes (Fig. 1B, fig. S1D). Within sCD3-lymphocytes, high expression of CD7 and CD103 identified the aberrant sCD3-lymphocytes in RCD2 individuals (referred to below as sCD3-CD103+) (*12*), which comprised 33% and 38% of CD45^+^ cells (Fig. 1, fig. S1F to H). These sCD3-CD103+ lymphocytes were also observed at 7.6% and 9.9% of CD45+ cells in two RCD1 individuals (RCD1-2 and RCD1-3) but were not discernible in the remaining RCD1 individuals nor in ANCD or celiac-negative controls (fig. S1H). RCD1 individuals had similar mixtures of B, T and epithelial cells to ANCD individuals, except RCD1-1 whose biopsy had an expansion of sCD3+ CD4-CD8-cells comprising 55% of total CD45^+^ cells (data S2, light blue in fig. S1D).

To test for lymphoma driver somatic mutations in each cell, custom scDNA-Seq amplicon panels targeted all the exons or focussed on mutation hotspots (data S2) in genes frequently mutated in RCD2 and EATL (*20*) including key genes of the JAK/STAT pathway (*JAK1*, *JAK3*, *STAT3, STAT5, SOCS1, SOCS3*), NF-κB signalling (*TNFAIP3, TNIP3*), epigenetic regulators (*DNMT3A*, *TET2* and *KMT2D*), and a regulator of mRNA translation (*DDX3X*). Somatic mutations were identified by filtering for nonsynonymous variants that clustered to a particular cell type (fig. S2A to D, see Methods), revealing 24 high confidence mutations in intestinal cells from 2/2 RCD2 individuals, 7/10 RCD1 individuals, 1/4 ANCD individuals, and 0/7 non-CD controls (table S2).

### Ongoing *TCR* recombination in aberrant sCD3-lymphocyte clones in RCD2

In RCD2 patients, scDNA-Seq revealed the sCD3-CD103+ lymphocytes had multiple driver mutations co-existing in the same cells. In individual RCD2-1, most sCD3-CD103+ cells (305/309 cells) comprised a single clone carrying four unambiguous driver mutations (Fig. 1C): *TNFAIP3^p.L324Qfs*6^* creating haploinsufficiency; hemizygous *DDX3X^p.T532A^* substituting an absolutely conserved threonine recurrently mutated in medulloblastoma and required for unwinding translationally-silenced RNA duplexes (*26*); *KMT2D^p.Q2380*^* preceding the catalytic SET methyltransferase domain; and *POT1^p.G236V^*.

In individual RCD2-2, the sCD3-CD103+ lymphocytes consisted of two clones with distinct driver mutations (Fig. 1C). A large clone (175/225 sCD3-CD103+ cells) carried *JAK1^p.G1097C^*, *TET2^p.N1118Kfs*11^*, and *KMT2D^p.C1408F^* substituting an absolutely conserved zinc-coordinating cysteine in the H3K4-methyl-binding fourth PHD finger where loss-of-function missense mutations are frequently found (*27, 28*). A smaller clone (22/225 sCD3-CD103+ cells) carried *JAK1^p.G1097D^* and *JAK3^p.M511I^*. Substitutions at Gly1097 in JAK1 occur in >40% of RCD2 individuals (*20*). Gly1097Asp has been demonstrated to be gain-of-function (GOF) (*29*) while Gly1097Cys is predicted GOF, as both introduce bulky side chains where the absence of a side-chain (Gly) is needed for binding of the inhibitor SOCS1 and the inhibitory ubiquitin ligase adaptor SOCS3 (*30*). The *JAK1* mutations in the two clones were called homozygous in almost every cell (Fig. 1C), apparently due to loss of the wild-type allele based on lower amplicon sequencing coverage across the *JAK1* locus in the sCD3-CD103+ cells (fig. S2E), and a high proportion of homozygous calls for heterozygous germline SNPs in the *JAK1* locus in single-cell RNA-sequencing of these cells (fig. S2F). Loss-of-heterozygosity (LOH) in chromosome 1p has previously been reported in RCD2 (*20*). To our knowledge this is the first evidence for two distinct sCD3-CD103+ clones with different driver mutations in an RCD2 individual, likely because resolving them requires mutation analysis at single cell resolution.

A complementary single cell multi-omics approach was employed to reveal whole transcriptome (scRNA-Seq) and full-length *TCR* mRNA sequences in conjunction with cell surface protein profiles (using oligo-barcoded antibodies) in thousands of single cells (Fig. 2A, data S3, see Methods). For the RCD2 individuals, UMAP plots of global mRNA and protein expression showed the aberrant sCD3-CD103+ lymphocytes formed a distinct population from other lymphocytes in the biopsies (fig. S3, A to B). At both mRNA and protein levels, sCD3-CD103+ lymphocytes differed extensively from sCD3+ CD8+ or CD4+ T cells present in the biopsies, although they shared with CD8 T cells expression of Fas ligand, perforin, granzyme b and expressed NK cell activating receptors *KLRK1*/NKG2D, *HCST* encoding NKG2D’s signalling partner DAP10, *NCR1*/NKP46, and *KLRC2* and *KLRD1* encoding the NKG2C-CD94 heterodimer (fig. S3, C to H). The sCD3-CD103+ lymphocytes expressed mRNAs encoding key transcription factors for ILC differentiation *NFIL3 (31, 32)* and *TOX (33)*, expressed *TBX21* (T-bet) which defines the ILC1 subset, but lacked mRNAs encoding transcription factors expressed in NK cells or other ILC subsets such as *EOMES*, *RORC* (ROR-γt), and *TCF7* (TCF-1) (*34*). The sCD3-CD103+ lymphocytes expressed high levels of *IKZF2* (Helios), similar to an ILC1 subset expressing rearranged TCRγ/δ mRNA (*35*) (fig. S3, I to J).

**Fig. 2.**
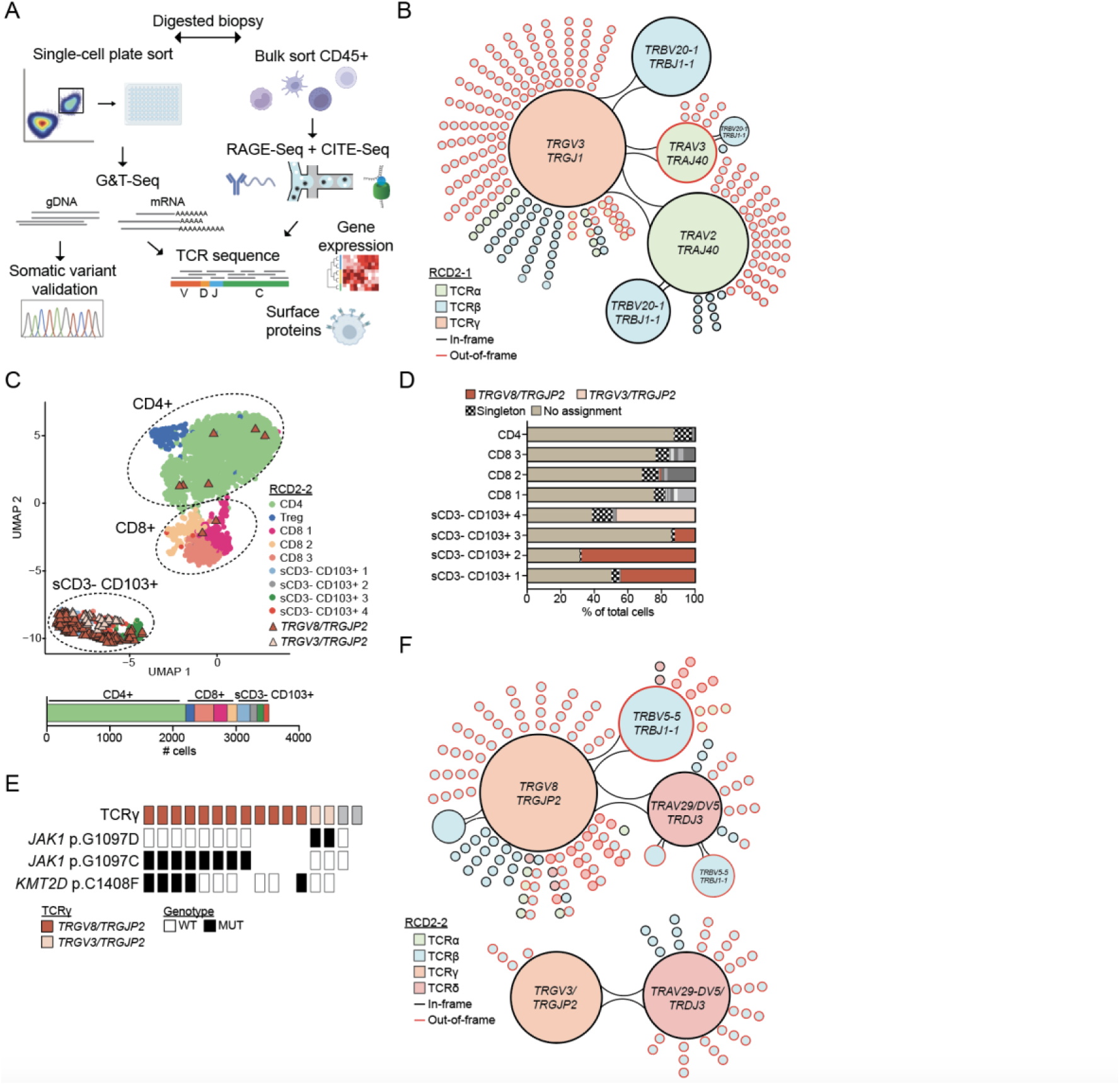
Intraclonal diversity of expressed TCR sequences in RCD2 individuals. **(A)** Strategy to match DNA mutations, TCR sequences, mRNA expression and cell surface protein profiles in single duodenal CD45+ cells. (**B**) Network plot depicting intraclonal diversity in sCD3-CD103+ cells from individual RCD2-1. Large subclones co-expressing the clonal TCRγ and recurring TCRβ or TCRα sequences denoted by connected large circles (n=433 cells also assigned TCRα and/or TCRβ chains). The smallest circles indicate one cell expressing a unique TCRβ and/or TCRα sequence but also expressing a recurrent TCR sequence denoted by the adjacent large circles. Clone sizes: *TRAV2/TRAJ40*, 188 cells; *TRAV2/TRAJ40* and *TRBV20-1/TRBJ1-1*, 27 cells; *TRBV20-1*/*TRBJ1-1*, 59 cells; *TRAV3/TRAJ40*, 22 cells. (**C)** UMAP of CD45+ sCD3+ and sCD3-CD103+ cells for individual RCD2-2 (n=3,528 cells total) using combined gene-expression and cell surface protein single cell data. The proportion of cell types are shown below the UMAP. Individual cells expressing clonal TCRγ chain sequences *TRGV8*/*TRGJP2* (n=189 cells) or *TRGV3*/*TRGJP2* (n=39 cells) are denoted by coloured triangles. (**D**) Assignment of unique TCRγ V(D)J chains to the cell clusters identified in the UMAP for individual RCD2-2. The *TRGV8*/*TRGJP2* and *TRGV3*/*TRGJP2* clonal sequences are highlighted in color as per the legend. Grey colours indicate TCRγ clones with size >1 cell. n=710 total cells assigned a TCRγ chain amongst 207 “CD3-CD103+ 1” cells, 109 “CD3-CD103+ 2” cells, 105 “CD3-CD103+ 3” cells, 83 “CD3-CD103+ 4” cells, 306 “CD8 1” cells, 212 “CD8 2” cells, 158 “CD8 3” cells, 2,209 CD4 cells. (**E**) G&T-Seq analysis of CD45+ sCD3-CD19-flow sorted duodenal cells from individual RCD2-2. For each individual cell, TCRγ chains reconstructed from mRNA gene expression were linked to *JAK1* and *KMT2D* mutations sequenced from DNA. A total of n=16 cells were identified with TCRγ chains, with colors representing the indicated clonotype, while TCRγ chains depicted in grey are clonally unrelated to *TRGV8*/*TRGJP2* and *TRGV3*/*TRGJP2.* (**F**) Network plot depicting intraclonal diversity of expressed TCRδ, TCRβ and TCRα sequences in sCD3-CD103+ cells co-expressing the clonal *TRGV8*/*TRGJP2* TCRγ chain (n=136 cells also assigned TCRα/β/δ) or the *TRGV3/TRGJP2* TCRγ chain (n=40 cells also assigned TCRα/β/δ) from individual RCD2-2. Clone sizes for *TRGV8/TRGJP2* TCRγ clone: *TRAV29/DV5*, 26 cells; *TRAV29/DV5* and *TRBV5-5/TRBJ1-1*, 6 cells; *TRBV5-5/TRBJ1-1*, 35 cells. Clone sizes for *TRGV3/TRGJP2* TCRγ clone: *TRAV29-DV5/TRDJ3*, 36 cells.

Single cell full-length TCR sequences reconstructed from scRNA-Seq (*36*) revealed an identical, in-frame rearranged TCRγ chain in most of the aberrant sCD3-CD103+ lymphocytes with an assignable TCRγ sequence within each clone. In individual RCD2-1, 99.5% of assignable cells expressed a sequence joining the *TRGV3* variable element to the *TRGJ1* joining element (fig. S4, A to B). However, there was remarkable intraclonal diversity with expression of different, mostly out-of-frame recombined *TCRβ VDJ* mRNA sequences in individual cells, comprising 216 distinctly recombined singletons out of 306 cells with assigned *TCRβ* sequence (Fig. 2B, fig. S4C to G). Larger subclones expressed an in-frame recombined *TRBV20-1/TRBJ1-1* TCRβ mRNA (75/306 cells), a subset of which co-expressed an in-frame recombined *TRAV2/TRAJ40* TCRα mRNA. Cells with this TCRα mRNA represented a dominant subclone (188/233 cells with assigned TCRα), while a smaller subclone associated with a different recombined TCRβ mRNA expressed a distinct, out-of-frame recombined *TRAV3/TRAJ40* mRNA (22/233 cells). Analysis of a biopsy collected four months prior revealed the pre-existing dominant TCRα and TCRβ subclones (fig. S4, H to J). The subset of cells with in-frame recombined TCRα and TCRβ mRNAs did not have discernibly increased expression of sCD3 (fig. S4K).

In individual RCD2-2 the aberrant sCD3-CD103+ lymphocytes comprised two clones expressing different in-frame recombined TCRγ chain mRNA sequences: 74% of sCD3-CD103+ cells with assigned TCRγ expressed *TRGV8* joined to *TRGJP2,* and 16% expressed *TRGV3* joined to *TRGJP2* (Fig. 2C to D). While both clones had similar expression of CD103, cytotoxic genes, and NK receptors (fig. S3, G to H), in UMAP of global mRNA and protein expression they formed separately resolved subclusters within the sCD3-CD103+ population (Fig. 2C, fig. S4L). To match TCRγ clonal rearrangements with driver mutations we performed plate-based G&T-Seq where mRNA and genomic DNA can be sequenced from the same single cell (*37*) (Fig. 2A). Cells expressing *TRGV8/TRGJP2* mRNA carried *JAK1^p.G1097C^* and *KMT2D^p.C1408F^*, whereas cells expressing *TRGV3/TRGJP2* mRNA carried *JAK1^p.G1097D^* (Fig. 2E).

The two RCD2-2 clones nevertheless expressed an identical in-frame recombined TCRδ mRNA comprising *TRAV29-DV5* joined to *TRDDδ2,* two copies of *TRDD3,* and *TRDJ3* (28/67 cells assigned TCRα or TCRδ chains) (Fig. 2F, fig. S4M). Because the two clones also shared apparent LOH of the *JAK1* locus they appear to have a common clonal origin, diverging after *JAK1* LOH and TCRδ rearrangement to acquire distinct driver mutations and TCRγ rearrangements. Extensive intraclonal diversity was observed in both clones, comprising numerous cells with different recombined TCRβ mRNAs, mainly out-of-frame singleton subclones along with a few expanded subclones such as an out-of-frame recombined *TRBV5-5/TRBJ1-1* mRNA (32/103 TCRβ chain assigned cells) (Fig. 2F, fig. S4, D to G, N). We confirmed by plate-based G&T-Seq the co-expression in individual cells of *TRBV5-5/TRBJ1-1* TCRβ, *TRAV29-DV5/TRDJ3* TCRδ and *TRGV8/TRGJP2* TCRγ mRNAs (data S4).

Collectively, single cell TCR mRNA sequencing revealed extraordinary intraclonal diversity from ongoing *TCR* recombination in the aberrant sCD3-lymphocyte clones infiltrating the intestine of RCD2 patients that is likely to be missed by diagnostic PCR tests performed on total DNA from biopsies.

### Mutated clones of sCD3-intestinal lymphocytes in RCD1

As noted above, single cell analysis revealed populations of aberrant sCD3+ CD103+ lymphocytes in 2 of 10 RCD1 patients, with frequencies falling well below the diagnostic threshold for RCD2 (*14*). In individual RCD1-3, sCD3-CD103+ lymphocytes comprised 9.89% (147/1486) of CD45+ cells (fig. S1H) and comprised two driver-mutated clones: 77/147 cells carried *TET2^p.H1036Lfs*9^* and *STAT3^p.Y657_K658insSY^*, whereas 41/147 cells carried *STAT3^p.D661V^* (Fig. 3A).

**Fig. 3.**
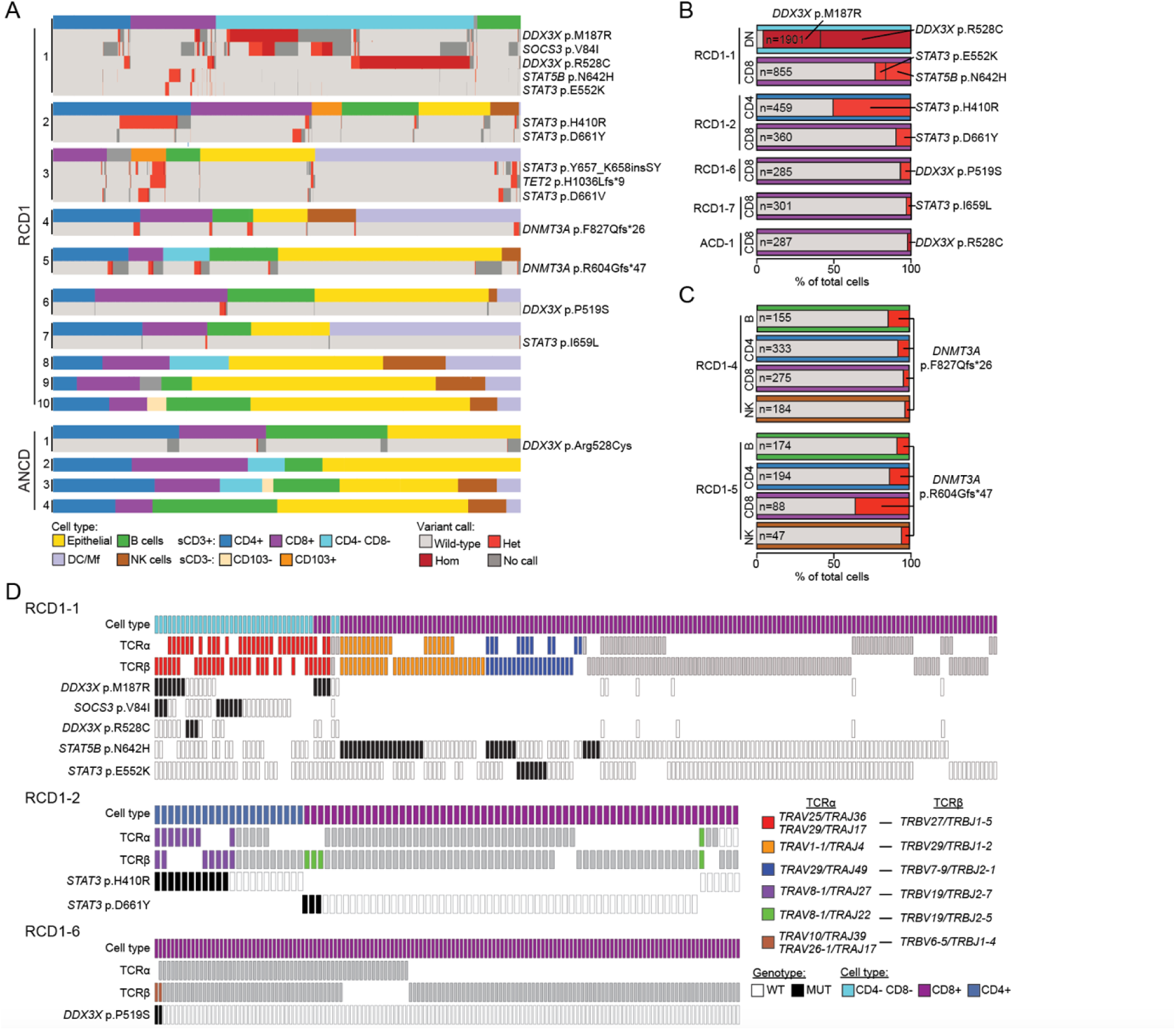
Single cell somatic DNA mutation detection in sCD3+ TCR+ T cells in RCD1 individuals. **(A)** Boxplots of 20,894 cells from 10 RCD1 individuals and 8,402 cells from 4 ANCD individuals assigning cell types and DNA mutation call by scDNA-Seq. (**B-C**) Frequencies of cells with the indicated mutations within their respective cell type (B), or if they were found in multiple cell types (C). Variant and cell type colors correspond to keys in (A). DN; CD4-CD8-. B; B cells. (**D**) G&T-Seq analysis of single CD45+ sCD3+ cells flow sorted as either CD4+, CD8+, or CD4-CD8-from individuals RCD1-1 (n=196 cells), RCD1-2 (n=110) and RCD1-6 (n=178). Cells are depicted if they had a TCRα and/or TCRβ V(D)J sequence determined. Clonal TCR sequences are color-coded, while cells with clonally unrelated TCRs are shown in grey. For each cell, indicated DNA mutation calls are shown below. Absence of a box indicates a cell with missing TCR or DNA mutation information.

### Expanded clones of sCD3+ TCR+ cells with driver mutations in RCD1

In contrast to the aberrant sCD3-CD103+ lymphocyte clones defining RCD2 individuals, putative driver mutations were found within clonally expanded sCD3+ T cells from duodenal biopsies in 6 of 10 RCD1 individuals (Fig. 3A, table S2). CD3 was expressed at comparable levels on the surface of mutation-bearing sCD3+ T cells and their wild-type counterparts in the same biopsy, in contrast to the aberrant sCD3-lymphocytes in RCD2 individuals (fig. S5A).

Individual RCD1-1, whose unusual expansion of intestinal sCD3+ CD4-CD8-T cells and resistance to steroid therapy led to chemotherapy with cladribine normally reserved for RCD2 patients (see supplementary text), had 4 different sCD3+ T cell clones with independent driver mutations. The large expansion of sCD3+ CD4-CD8-T cells consisted of two clones distinguished by acquisition of *DDX3X^p.R528C^* (1154/2511 cells) or *DDX3X^p.M187R^* (951/2511 cells) hemizygous mutations. The *DDX3X^p.M187R^*-bearing clone also carried *SOCS3^p.V84I^*. Additionally, two clones of sCD3+ CD8+ T cells carried mutations: *STAT5B^p.N642H^* in 147/832 cells, and a separate clone comprising 66/832 cells with *STAT3^p.E552K^* (Fig. 3B).

Two mutant sCD3+ T cell clones were found in individual RCD1-2. A large clone comprising 232/538 of sCD3+ CD4+ T cells carried *STAT3^p.H410R^*, while 37/470 sCD3+ CD8+ T cells had *STAT3^p.D611Y^* (Fig. 3B).

Single clones of mutated sCD3+ CD8+ T cells were found in RCD1-6 and RCD1-7 (Fig. 3B). In individual RCD1-6, 19/318 sCD3+ CD8+ T cells carried *DDX3X^p.P519S^*. In individual RCD1-7, 10/310 sCD3+ CD8+ T cells carried *STAT3^p.I659L^*.

Small nucleotide deletions in *DNMT3A* resulting in frameshifts and premature stop codons were identified in individuals RCD1-4 and RCD1-5 (Fig. 3C). In RCD1-4, 24/333 sCD3+ CD4+ T cells, 10/275 sCD3+ CD8+ T cells, 21/155 B cells, and 5/184 NK cells carried *DNMT3A^p.F827Qfs*26^*. In RCD1-5, 20/194 sCD3+ CD4+ T cells, 26/88 sCD3+ CD8+ T cells, 10/174 B cells, and 2/47 NK cells carried *DNMT3A^p.R604Gfs*47^*. The presence of these mutations in multiple lymphocyte subsets but not in epithelial cells (Fig. 3A) is consistent with acquisition in a hematopoietic stem or progenitor cell as occurs for *DNMT3A* loss-of-function mutations in clonal haematopoiesis.

None of the control patients whose biopsy proved to be negative for active CD had mutated sCD3+ T cells or sCD3-lymphocytes (Fig. 1C), but a single *DDX3X^p.R528C^* mutated sCD3+ T cell clone was found in individual ANCD-1 comprising 6/245 sCD3+ CD8+ T cells, lower than the frequency of mutant sCD3+ T cells in RCD1 individuals (Fig. 3B).

To compare the sensitivity of scDNA-Seq to previous methods of bulk targeted capture sequencing of total biopsies (*20, 21*), pooled cell DNA obtained from the same RCD1 and RCD2 biopsy samples used for scDNA-Seq was analysed by targeted capture sequencing of all exons in 73 genes recurrently mutated in lymphoma (data S5). Of 8 mutations identified in sCD3+ T cells by scDNA-Seq in the RCD1 patients, only 4 were detected by bulk targeted sequencing (fig. S5B). *DDX3X^p.M187R^*, *DDX3X^p.R528C^*, *STAT3^p.H410R^* and *DNMT3A^p.R604Gfs*47^* - present in 9–32% of total cells from scDNA-Seq - were identified by bulk targeted sequencing in 3-13% of total reads. Despite unique read depths ranging from 344-1208 reads across the relevant nucleotides, *STAT3^p.D661Y^*(8.1% of total cells), *STAT3^p.E552K^* (1.5% of total cells), *STAT5b^p.N642H^* (3.3% of total cells), and *DDX3X^p.P519S^* (1.7% of total cells) were below the limit of detection of the variant caller in the bulk targeted sequencing as they were present in <5 reads. Lower sensitivity may explain why driver mutations were not detected by bulk targeted sequencing in biopsies from 18 RCD1 individuals and 7 active CD individuals (*20*).

In individuals RCD1-1, RCD1-2, and RCD1-6 we matched individual driver mutations with recombined *TCR* mRNA sequences by sequencing targeted exons in genomic DNA and *TCR* mRNAs from individual sorted cells (Fig. 2A, G&T-Seq). This independent validation confirmed all the driver mutations identified by scDNA-Seq and revealed TCRαβ VDJ sequences belonging to each mutated clone (Fig. 3D, table S3, data S4). In RCD1-1, the sCD3+ CD4-CD8-T cells co-expressed an identical, in-frame recombined *TRBV27/TRBJ1-5* TCRβ mRNA sequence and two different in-frame recombined TCRα mRNAs employing *TRAV25/TRAJ36* or *TRAV29-DV5/TRAJ17* (Fig. 3D). Different cells within this large clone carried either *DDX3X^p.M187R^*or *DDX3X^p.R528C^*, indicating that two subclonal branches descended from a single T cell and independently acquired different *DDX3X* mutations before becoming co-dominant progeny. A subset of both subclonal branches expressed CD8 albeit at lower levels than other CD8 cells in the sample (fig. S5C).

In the same individual RCD1-1, *STAT5B^p.N642H^* was found in different T cells from the *DDX3X* mutations and limited to sCD3+ CD8+ T cells (Fig. 3D). Of 28 *STAT5B^p.N642H^* mutant cells, 18 expressed *TRBV29-1/TRBJ1-2* mRNA (denoted orange in Fig. 3D) whereas 7 expressed *TRBV7-9/TRBJ2-1* (denoted blue in Fig. 3D). The *TRBV7-9/TRBJ2-1* CD8+ clone also contained a subset of cells with *STAT3^p.E552K^*albeit different cells from those with *STAT5B^p.N642H^* (Fig. 3D, fig. S5D). No cells were found with both mutations either in the G&T-Seq analysis here or in the scDNA-Seq analysis above, consistent with independent acquisition of *STAT5B^p.N642H^*in the two T cell clones and acquisition of *STAT3^p.E552K^* in a separate branch of the *TRBV7-9* clone.

In individual RCD1-2, *STAT3^p.H410R^* sCD3+ CD4+ T cells expressed in frame recombined *TRBV19/TRBJ2-7* and *TRAV8-1/TRAJ27* mRNAs, whereas *STAT3^p.D611Y^* sCD3+ CD8+ T cells expressed in-frame *TRBV19/TRBJ2-5* and *TRAV8/TRAJ22* mRNAs (Fig. 3D). In individual RCD1-6, *DDX3X^p.P519S^* sCD3+ CD8+ T cells expressed in frame *TRBV6-5/TRBJ1-4* and *TRAV10/TRAJ39 & TRAV26-1/TRAJ17* mRNAs (Fig. 3D).

None of the TCRαβ sequences carried by the mutated T cell clones expressed semi-invariant TCR sequences found on mucosal-associated invariant T (MAIT) cells (*TRAV1-2-TRAJ33/12/20*) (*38*) or invariant natural killer T (iNKT) cells (*TRAV10-TRAJ18*) (*39*) suggesting the T cell clones with driver mutations are distinct from these unconventional T cell populations (table S3).

### Characterisation of driver mutations found in sCD3+ clones

The STAT3 mutations were concentrated in the Src homology 2 (SH2 domain) that mediates dimerization and activation of STAT proteins, corresponding to recurrent GOF mutations in RCD2, EATL, and T cell large granular lymphocyte (T-LGL) leukemia (*20, 21, 40-42*) (Fig. 4A). Consistent with previous evidence that p.D661Y and p.H410R in the DNA binding domain increase STAT3 phosphorylation and transcriptional activity (*20, 43, 44*), these mutations, as well as p.E552K in the linker domain, increased STAT3 transcriptional activity in a luciferase reporter assay (Fig. 4B), increased phosphorylation of Tyr705 in STAT3, and delayed dephosphorylation (fig. S6, A to B). Mice with targeted STAT3 GOF mutations have exaggerated clonal expansions of CD8 effector T cells (*45, 46*).

**Fig. 4.**
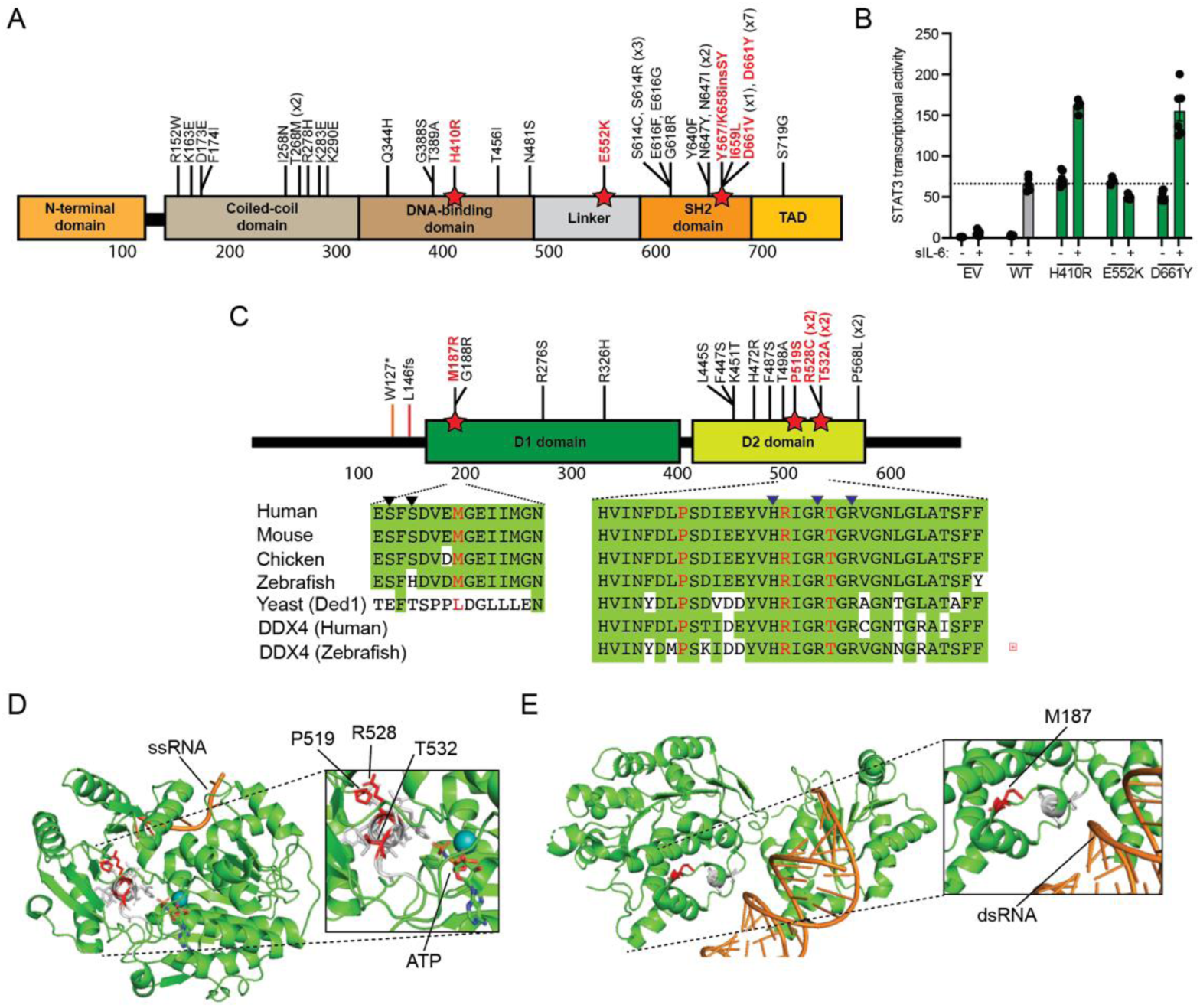
Characterisation of somatic mutations in sCD3+ T cell clones. **(A)** STAT3 protein domains and location of the 39 somatic mutations reported in RCD2 and EATL. Mutations in red are those identified in this study. SH2, Src Homology 2. TAD, transactivating domain. (**B**) Luciferase assay on STAT3^−/-^ A4 cells transfected with empty vector (EV), wild-type *STAT3* (WT), or the indicated *STAT*3 variants, together with the pGL4.47 luciferase reporter construct and an expression vector for *Renilla* luciferase. After 24 h, transfected cells were left untreated (“-“) or treated (“+”) with 100 ng/ml sIL-6 for 24 h. Mean luciferase activity normalized against the luminescence in unstimulated EV transfected cells and SEM of n=6 samples per condition. Dashed line: mean luciferase for WT transfected cells. (**C**) DDX3X protein domains and location of 19 somatic mutations reported in RCD2 and EATL. Mutations in red are those identified in this study. Amino acid alignments for human DDX3X amino acids 180-194 and 512-543 in the indicated species and other DEAD-Box helicases. Black triangles: RNA contact residues; blue triangles: ATP contact residues. (**D**) Structure of *Drosophila* Vasa/DDX4 (PDB:2DB3) post-unwound state bound to ATP and single stranded (ss) RNA. Residues corresponding to human DDX3X Pro519, Arg528 and Thr532 are shown in red. (**E**) Structure of human DDX3X (PBD:5O5F) pre-unwound state bound to double stranded (ds) RNA. M187 residue is highlighted.

The *DDX3X* mutations correspond to recurring mutations in RCD2, EATL and B cell lymphoma (*47*) and substitute absolutely conserved amino acids within the RNA helicase D1 and D2 domains (*48*) (Fig. 4C). Structural studies of human DDX3X and the *Drosophila* paralogue Vasa/DDX4 show the D2 domain rotates 180° between the pre-unwound state bound to dsRNA in the absence of ATP and the post-unwound state bound to ssRNA and nonhydrolyzable ATP analogue (*49, 50*). The DDX3X mutation hotspot helix between Pro519 and Arg534 contributes key side-chains directly binding ATP (*49*) (Fig. 4D). Substitution of Thr532 in human DDX3X abolishes helicase and ATPase activity (*48*). The *DDX3X^p.M187R^* mutation changes a buried hydrophobic residue to a polar residue in a loop that binds dsRNA in the pre-unwound structure (*50*) (Fig. 4E).

*STAT5B^p.N642H^* is an unambiguous driver mutation in the SH2 domain found frequently in γδ T cell-derived lymphoma, NK/T cell lymphoma, LGL, NKTL, T cell acute lymphoblastic leukemia, and monomorphic epitheliotropic intestinal T-cell lymphoma (MEITL; previously classified as EATL type II) (*51–55*). It enhances phospho-STAT5B dimerization to create a strong GOF driving increased expression of target genes (*53, 56*) and driving CD8^+^ effector memory T-cell lymphoma/leukemia in transgenic mice (*56–58*).

The *DNMT3A* frameshift mutations truncate the DNA methyltransferase domain to create unambiguous loss-of-function alleles (*59*) (fig. S6C).

The somatic mutations acquired by T cells accumulating in the intestine of RCD1 patients are thus skewed in favour of functional alterations that parallel driver mutations in T cell neoplasia, RCD2 and EATL.

### Larger clonal expansions and inflammatory gene expression programs in mutated T cell clones

In the three RCD1 individuals where clonal TCRαβ mRNA sequences had been determined and could be used as markers of particular driver-mutated sCD3+ T cells, we next investigated how the mutated clones compared to other T cell clones in the same biopsy by simultaneous measurement of TCR mRNA sequence, global mRNA expression, and expression of 159 cell surface protein markers (*36, 60*) (Fig. 5, fig. S7-S10).

**Fig. 5.**
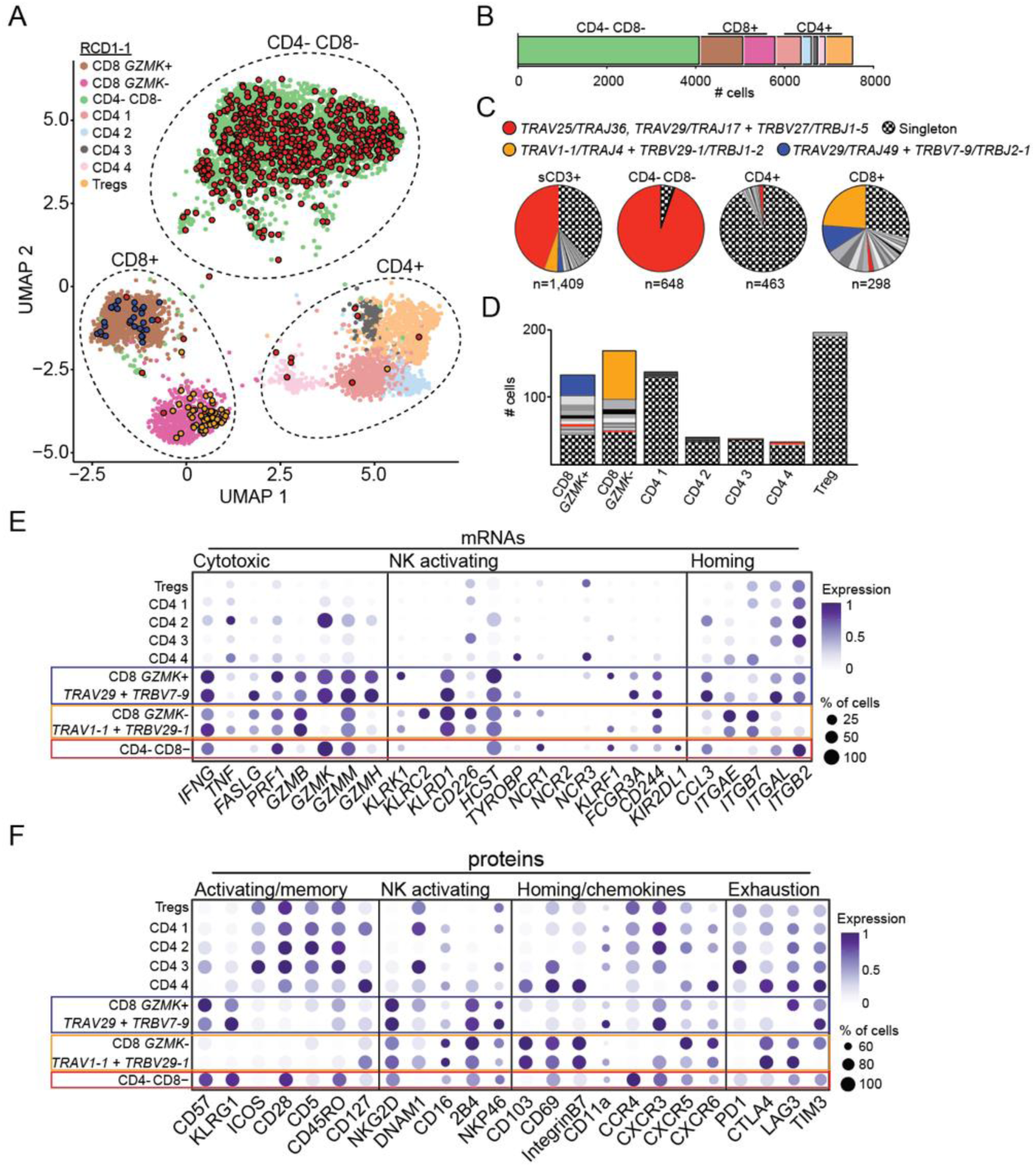
Gene expression programs and clonal expansions of mutated T cell clones in RCD1-1. **(A)** UMAP of CD45+ sCD3+ cells from patient RCD1-1 (n=7,569 cells) using combined gene-expression and cell surface protein data. Clusters are denoted by colours and labelled by cell type. Cells expressing clonal TCRαβ sequences denoted in (C) are highlighted in the UMAP. **(B)** Relative proportions of cell types identified by UMAP. (**C-D**) Proportion (C) and number of cells (D) in the indicated cell types assigned paired TCRα and TCRβ sequences (n=2,818 cells) corresponding to clones with driver mutations (color coded). Other expanded clones (grey) and unique singletons (pattern) are also shown. (**E-F**) Dot plots showing relative expression of selected mRNAs (E) and proteins (F) in the indicated cell type. Colored boxes mark cell subsets carrying driver mutations. The size of each dot represents the percentage of cells with non-zero expression, and colour intensity represents scaled log expression averaged over all cells.

In individual RCD1-1, UMAP analysis of global mRNA expression coupled with protein expression separated the intestinal sCD3+ T cells into three major subsets: CD4-CD8-(54.4%), CD4+ CD8-(23.1%), and two well separated subsets of CD4-CD8+ cells (collectively 22.6%). (Fig. 5, A to B, fig. S7, A to D). T cells expressing clonal TCR mRNA sequences marking *DDX3X* mutant cells were almost exclusively in the CD4-CD8-Cluster, comprised the majority of cells with an assigned TCR sequence in this cluster, and were the largest clone of T cells among all CD3+ cells in the biopsy (Fig. 5A, C).

In the two CD8+ cell subsets of RCD1-1, one expressed high *GZMK* mRNA and little CD103 protein or its mRNAs *ITGAE* and *ITGB7*, while the other lacked *GZMK* but expressed high CD103 protein and mRNAs and higher *GZMB*, corresponding to two CD8 effector T cell subsets in rheumatoid arthritis synovium and ulcerative colitis (*61*) and in healthy and transplanted intestine (*62*). A detailed comparison of gene and protein expression between these subsets is provided in fig. S7, E to I, but for simplicity we refer to these subsets here as CD8+ *GZMK*+ and CD8+ *GZMK*-. Cells expressing TCR mRNAs marking the clone with *STAT5B^p.N642H^* clustered in the CD8+ *GZMK*-subset (Fig. 5A), which was three times larger than the next largest T cell clone in this subset (Fig. 5D). The third largest T cell clone in the biopsy comprised cells expressing TCR mRNAs marking the clone with branches bearing *STAT3^p.E552K^* or *STAT5B^p.N642H^* mutations. This clone clustered in the CD8+ *GZMK*+ subset (Fig. 5A) where it was two times larger than the next largest T cell clone in that subset (Fig. 5D).

The finding that the three clones carrying driver mutations corresponded to the three most expanded clones of T cells in the biopsy is consistent with these mutations conferring a growth or survival advantage. While the three mutated T cell clones segregated into T cell subsets with globally different mRNA and protein expression, they nevertheless shared strong expression of mRNAs encoding inflammatory or cytotoxic proteins interferon gamma, perforin, a range of granzymes, and NK-cell activating receptor subunits NKG2D, DAP10 and 2B4 (Fig. 5, E to F).

We analysed additional duodenal biopsies of individual RCD1-1 taken 6 and 20 months following the initial sample (fig. S8A). The individual had previously received cladribine treatment prior to the initial sample and has remained well although has worsening enteropathy (table S1). Both the *DDX3X* mutant sCD3+ CD4-CD8-Clones and the *STAT5* and *STAT3* mutant sCD3+ CD8+ clones, defined by *TCR* sequence, were detected at both additional timepoints and remained the top three most frequent sCD3+ clones, and the most frequent clone within their respective cell type (fig. S8B to F). The mutated clones retained their cytotoxic gene and protein expression profile, with consistent expression of cytotoxic and inflammatory markers (fig. S8, G to H). scDNA-Seq confirmed the persistence of the clones at the DNA mutation level and showed that within the sCD3+ CD4-CD8-Clone, the *DDX3X^p.M187R^ SOCS3^p.V84I^* clonal branch increased in proportion relevant to the *DDX3X^p.R528C^* branch (fig. S8, I to J), potentially reflecting additional growth advantage provided by two driver mutations.

Analysis of individuals RCD1-2 and RCD1-6 yielded similar findings (see fig. S9, S10). In individual RCD1-2 the cells expressing *TCR* mRNA marking the *STAT3^p.H410R^* CD4+ T cell clone represented the largest T cell clone in the biopsy, comprising 32% of CD3+ cells (fig. S9, A to D). This CD4+ clone formed two separate clusters in UMAP analysis of overall gene and protein expression (fig. S9B), one being CD103+ *GZMK-* and the other CD103-*GZMK*+, but both subsets strongly expressing *IFNG* and expressing *TNF*, *GZMB* and a range of NK-activating receptors (fig. S9, E to J). CD8+ T cells also formed two distinct clusters in UMAP analysis corresponding to CD103+ *GZMK-* and CD103-*GZMK*+ subsets (fig. S9B, K). Cells with TCR mRNAs marking the *STAT3^p.D661Y^* CD8+ T cell clone represented the largest clone among sCD3+ CD8+ T cells (11.7% of CD8 cells) and were exclusively within the *GZMK*+ subset where they accounted for 20% of that subset (fig. S9D) and expressed higher levels of *IFNG, FASLG, PRF1* and *GZMB* than the other T cell clones contributing to this subset (fig. S9, E to F, L to N).

In RCD1-6 three clusters of sCD3+ CD8+ cells were identified in UMAP analysis of global gene and protein expression, two of which were CD103+ *GZMK*-Cells (*GZMK*-1 and *GZMK*-2), and the other CD103-*GZMK*+ cells (fig. S10A). Cells expressing *TCR* mRNAs marking the *DDX3X^p.P519S^*sCD3+ CD8+ clone represented the second most frequent TCRαβ clone amongst total sCD3^+^ cells (6.2%) and sCD3+ CD8^+^ cells (7.7%) and were exclusively within the two clusters of *GZMK*-Cells (fig. S10B). However, the driver mutated T cells differed from other CD8 cells in each cluster by strong expression of *GZMK* and other cytotoxic/inflammatory mRNAs, and lower expression of CD103 and *ITGB7* mRNA (fig. S10 C to I).

In summary, these results demonstrate that in RCD1, large expansions of driver-mutated sCD3+ T cell clones accumulate and persist over time, and express inflammatory and cytotoxic gene and protein profiles that may contribute to ongoing enteropathy.

## Discussion

Celiac disease that fails to respond to strict dietary gluten exclusion presents a major clinical problem as persistent enteropathy and symptoms are linked to increased morbidity and reduced quality of life. By leveraging the sensitivity of new single cell multiomic technologies, we revealed lymphoma driver gene mutations in intestinal sCD3+ T cell clones or sCD3-ILC/T cell progenitor clones expressing a range of inflammatory and cytotoxic mRNAs in 7 of 10 people with RCD1. Mutated intestinal lymphocytes may account for non-responsive CD in a much larger proportion of patients than previously recognised. While an understanding of RCD2 pathogenesis based on aberrant sCD3-lymphocytes has enabled novel therapies to be tested and trialled, such as anti-IL-15 antibodies (*63*) and JAK inhibitors (*64, 65*), to date there have been no breakthroughs in understanding RCD1 development. In RCD1 systemic corticosteroids, immunomodulators, and biologics have been suggested in attempts to control inflammation and symptoms, but their long-term use is associated with significant side effects and there is limited data to guide appropriate treatment choices (*66*). Studies to better understand RCD have also been hamstrung by the lack of biomarkers to differentiate true RCD1 that occurs in the absence of gluten from active CD caused by trace or inadvertent gluten exposure.

Many consider RCD1 to simply occur in a subset of patients who are immunologically ultra-sensitive to low level gluten exposure. Supporting this notion is an HLA gene dose effect where HLA-DQ2.5 homozygosity enables presentation of 4x as many gluten peptides and markedly increases the risk of CD (*67*) and is found in 21% of uncomplicated CD patients compared to 44-67% of patients with RCD2, although there was no significant increase in RCD1 patients (*25, 68*). The somatic mutations in large clones of infiltrating sCD3+ T cells or sCD3-progenitors found here in RCD1 may have a similar magnifying effect.

Although the current view of RCD2 is a discrete clinicopathologic entity, our findings support the notion that acquisition of somatic mutations separate to the ILC/progenitor T cell compartment may be important in the development of self-sustaining disease. It suggests a model of celiac disease where clinical phenotypes exist on a spectrum: no intestinal lymphocyte mutations (well treated disease); rare, mutated sCD3+ T cell clones (active disease); frequent, large mutated sCD3+ T cell or sCD3-ILC/progenitor T cell clones (RCD1); and very large, highly mutated, aberrant sCD3-Clones (RCD2).

The discovery here that the aberrant sCD3-CD103+ lymphocyte clones in RCD2 are undergoing extensive ongoing *TCR* recombination may explain progression of 60-80% of RCD2 individuals to aggressive EATL within 5 years (*23–25*). ILCs and sCD3+ T cells share progenitor cells in the fetal thymus with the potential to differentiate into NK cells, other ILC subsets, or sCD3+ TCRγδ+ or TCRαβ+ cells (*19*). These progenitors seed the intestine where they retain the capacity for NOTCH and IL-15 induced differentiation into NK cells and sCD3+ TCRαβ+ cells in uncomplicated CD but appear blocked in differentiation in RCD2 (*15, 18*). Through RAG1/RAG2-mediated off-target chromosomal breaking and joining, pre-leukemic progenitors of acute lymphoblastic leukemia acquire many of the driver mutations found in these aggressive leukemias (*69, 70*). These off-target structural variants cannot be readily identified by current single cell DNA sequencing methods. By scRNA sequencing there was no evidence for ongoing *TCR* recombination in the driver-mutated sCD3+ T cell clones in the RCD1 patients. Absence of RAG-mediated mutagenesis in mature T cells may explain why similar driver mutations such as *STAT3* GOF promote exaggerated T cell clonal expansion but rare progression to aggressive leukemias or lymphomas (*45, 46*).

Two explanations can be considered for the mutated sCD3+ T cells or sCD3-lymphocytes forming larger clones than other similar T cells in the biopsies. First, the driver mutations confer a growth or survival advantage to these cells. Alternatively, mutations in the 12 genes tested – and up to 53 in some individuals - may simply be more likely to arise in T cells that have undergone more cell divisions. Several observations here strongly favour the first possibility. Foremost is that the mutations are concentrated in structural parts of the encoded proteins that create GOF in the case of *STAT3* and *STAT5*, or loss-of-function of *DDX3X*. *STAT3* GOF mutations drive accumulation of larger clones of sCD3+ CD8 effector T cells to precipitate autoimmune disease in mice (*45, 46*). Second, there were striking examples of convergent acquisition of different loss-of-function *DDX3X* mutations in different branches of the same T cell clone, or convergent acquisition of the same *STAT5* GOF mutation in different T cell clones within the same individual.

A limitation of our study is that we have only tested mutations in a small proportion of lymphoma driver genes and are likely to underestimate the presence of sCD3+ T cell or sCD3-CD103+ lymphocyte clones with driver mutations. Another limitation is that we cannot resolve whether the presence of these mutated clones is a cause or simply a consequence of chronic inflammation associated with RCD. Chronic intestinal inflammation and proliferation of sCD3+ T cells and progenitor cells is likely to increase the probability these cells acquire a somatic mutation that provides it with a growth or survival advantage. While these clones were found in 7/10 RCD1 individuals compared to 1/4 newly diagnosed active CD individuals, and the clones were more numerous in the RCD1 individuals, future prospective studies with large numbers of newly diagnosed individuals will be needed to determine the extent to which the presence of mutated T cell clones predicts non-response to gluten-free diet.

Many autoimmune diseases are hypothesized to have an external trigger that initiates a self-limiting phase of T cell activation and autoantibody production, followed later by transition to a chronic relapsing or sustained, more damaging phase. The findings here support the general concept that the self-sustained phase of autoimmune diseases may reflect acquisition of lymphoma driver mutations in T cells or ILCs resulting in their accumulation to higher numbers and greater pathological effect. Identifying a threshold frequency of mutated T cells or ILCs in CD may represent a positive biomarker informing better treatments for non-responsive CD.

## Data Availability

All data produced in the present study are available upon reasonable request to the authors

## Acknowledgements

We are grateful to all the volunteers who participated in this study. We thank the members of the Goodnow laboratory for helpful discussions. We thank the Garvan-Weizmann Centre for Cellullar Genomics, the Kinghorn Cancer Centre for Clinical Genomics, the Garvan Sequencing Platform and the Ramaciotti Centre UNSW Sydney for technical services. We thank Lee Henneken for help with sample collection.

## Author contributions

Conceptualization: MS, CCG, FL

Data curation: RHYL, JS, MAF

Formal analysis: RHYL, JS, MAF

Funding acquisition: MS, CCG, FL, DS

Investigation: MS, CM, TA, JM, ER, MF, MYH, PB,

Methodology: MS, CCG, FL

Project administration: MS, CCG, FL

Resources: JAT, GA, RC, AP, MVL, ADS, DS, SR, MYH, CSM, SGT, CCG, FL

Software: RHYL, JS, MAF, KJLJ, AC, TGA, IWD

Supervision: MS, CCG, FL

Visualization: MS, RHYL, JS, MAF

Writing – original draft: MS, CCG, FL

Writing – review and editing: MS, CCG, FL, JAT, DS

## Supplementary Materials

Materials and Methods

Supplementary Text

Figs. S1 to S10

Tables S1 to S3

References (#71-98)

Data S1 to S8

## References and Notes

1. P. Singh et al., Global Prevalence of Celiac Disease: Systematic Review and Meta-analysis. Clin Gastroenterol Hepatol 16, 823–836 e822 (2018).

2. R. Iversen, L. M. Sollid, The Immunobiology and Pathogenesis of Celiac Disease. Annu Rev Pathol 18, 47–70 (2023).

3. M. N. Marsh, Gluten, major histocompatibility complex, and the small intestine. A molecular and immunobiologic approach to the spectrum of gluten sensitivity (’celiac sprue’). Gastroenterology 102, 330–354 (1992).

4. A. Levescot, G. Malamut, N. Cerf-Bensussan, Immunopathogenesis and environmental triggers in coeliac disease. Gut 71, 2337–2349 (2022).

5. B. Jabri et al., Selective expansion of intraepithelial lymphocytes expressing the HLA-E-specific natural killer receptor CD94 in celiac disease. Gastroenterology 118, 867–879 (2000).

6. A. Kornberg, et al., Gluten induces rapid reprogramming of natural memory αβ and γδ intraepithelial T cells to induce cytotoxicity in celiac disease. Sci Immunol 8, eadf4312 (2023).

7. B. Meresse et al., Coordinated induction by IL15 of a TCR-independent NKG2D signaling pathway converts CTL into lymphokine-activated killer cells in celiac disease. Immunity 21, 357–366 (2004).

8. B. Meresse et al., Reprogramming of CTLs into natural killer-like cells in celiac disease. J Exp Med 203, 1343–1355 (2006).

9. H. A. Penny et al., Non-Responsive and Refractory Coeliac Disease: Experience from the NHS England National Centre. Nutrients 14, (2022).

10. J. A. Tye-Din, Review article: Follow-up of coeliac disease. Aliment Pharmacol Ther 56 Suppl 1, S49–S63 (2022).

11. J. F. Ludvigsson et al., The Oslo definitions for coeliac disease and related terms. Gut 62, 43–52 (2013).

12. C. Cellier et al., Abnormal intestinal intraepithelial lymphocytes in refractory sprue. Gastroenterology 114, 471–481 (1998).

13. A. Rubio-Tapia, J. A. Murray, Classification and management of refractory coeliac disease. Gut 59, 547–557 (2010).

14. W. H. Verbeek et al., Flow cytometric determination of aberrant intra-epithelial lymphocytes predicts T-cell lymphoma development more accurately than T-cell clonality analysis in Refractory Celiac Disease. Clin Immunol 126, 48–56 (2008).

15. J. Ettersperger et al., Interleukin-15-Dependent T-Cell-like Innate Intraepithelial Lymphocytes Develop in the Intestine and Transform into Lymphomas in Celiac Disease. Immunity 45, 610–625 (2016).

16. G. J. Tack et al., Origin and immunophenotype of aberrant IEL in RCDII patients. Mol Immunol 50, 262–270 (2012).

17. J. M. Tjon et al., Defective synthesis or association of T-cell receptor chains underlies loss of surface T-cell receptor-CD3 expression in enteropathy-associated T-cell lymphoma. Blood 112, 5103–5110 (2008).

18. F. Schmitz et al., Identification of a potential physiological precursor of aberrant cells in refractory coeliac disease type II. Gut 62, 509–519 (2013).

19. S. B. Shin, K. M. McNagny, ILC-You in the Thymus: A Fresh Look at Innate Lymphoid Cell Development. Front Immunol 12, 681110 (2021).

20. S. Cording et al., Oncogenetic landscape of lymphomagenesis in coeliac disease. Gut 71, 497–508 (2022).

21. C. R. Soderquist et al., Immunophenotypic Spectrum and Genomic Landscape of Refractory Celiac Disease Type II. Am J Surg Pathol 45, 905–916 (2021).

22. C. Cellier et al., Refractory sprue, coeliac disease, and enteropathy-associated T-cell lymphoma. French Coeliac Disease Study Group. Lancet 356, 203–208 (2000).

23. A. Al-Toma, W. H. Verbeek, M. Hadithi, B. M. von Blomberg, C. J. Mulder, Survival in refractory coeliac disease and enteropathy-associated T-cell lymphoma: retrospective evaluation of single-centre experience. Gut 56, 1373–1378 (2007).

24. S. Daum et al., High rates of complications and substantial mortality in both types of refractory sprue. Eur J Gastroenterol Hepatol 21, 66–70 (2009).

25. G. Malamut et al., Presentation and long-term follow-up of refractory celiac disease: comparison of type I with type II. Gastroenterology 136, 81–90 (2009).

26. A. L. Lennox et al., Pathogenic DDX3X Mutations Impair RNA Metabolism and Neurogenesis during Fetal Cortical Development. Neuron 106, 404–420 e408 (2020).

27. V. Faundes, G. Malone, W. G. Newman, S. Banka, A comparative analysis of KMT2D missense variants in Kabuki syndrome, cancers and the general population. J Hum Genet 64, 161–170 (2019).

28. A. M. Stroynowska-Czerwinska et al., Clustered PHD domains in KMT2/MLL proteins are attracted by H3K4me3 and H3 acetylation-rich active promoters and enhancers. Cell Mol Life Sci 80, 23 (2023).

29. R. Crescenzo et al., Convergent mutations and kinase fusions lead to oncogenic STAT3 activation in anaplastic large cell lymphoma. Cancer Cell 27, 516–532 (2015).

30. N. P. D. Liau et al., The molecular basis of JAK/STAT inhibition by SOCS1. Nat Commun 9, 1558 (2018).

31. T. L. Geiger et al., Nfil3 is crucial for development of innate lymphoid cells and host protection against intestinal pathogens. J Exp Med 211, 1723–1731 (2014).

32. C. Seillet et al., Nfil3 is required for the development of all innate lymphoid cell subsets. J Exp Med 211, 1733–1740 (2014).

33. C. R. Seehus et al., The development of innate lymphoid cells requires TOX-dependent generation of a common innate lymphoid cell progenitor. Nat Immunol 16, 599–608 (2015).

34. C. Harly et al., The transcription factor TCF-1 enforces commitment to the innate lymphoid cell lineage. Nat Immunol 20, 1150–1160 (2019).

35. L. Mazzurana et al., Tissue-specific transcriptional imprinting and heterogeneity in human innate lymphoid cells revealed by full-length single-cell RNA-sequencing. Cell Res 31, 554–568 (2021).

36. M. Singh et al., High-throughput targeted long-read single cell sequencing reveals the clonal and transcriptional landscape of lymphocytes. Nat Commun 10, 3120 (2019).

37. I. C. Macaulay et al., G&T-seq: parallel sequencing of single-cell genomes and transcriptomes. Nat Methods 12, 519–522 (2015).

38. R. Reantragoon et al., Antigen-loaded MR1 tetramers define T cell receptor heterogeneity in mucosal-associated invariant T cells. J Exp Med 210, 2305–2320 (2013).

39. O. Lantz, A. Bendelac, An invariant T cell receptor alpha chain is used by a unique subset of major histocompatibility complex class I-specific CD4+ and CD4-8-T cells in mice and humans. J Exp Med 180, 1097–1106 (1994).

40. H. L. Koskela et al., Somatic STAT3 mutations in large granular lymphocytic leukemia. N Engl J Med 366, 1905–1913 (2012).

41. A. B. Moffitt et al., Enteropathy-associated T cell lymphoma subtypes are characterized by loss of function of SETD2. J Exp Med 214, 1371–1386 (2017).

42. A. Nicolae et al., Mutations in the JAK/STAT and RAS signaling pathways are common in intestinal T-cell lymphomas. Leukemia 30, 2245–2247 (2016).

43. E. Andersson et al., Activating somatic mutations outside the SH2-domain of STAT3 in LGL leukemia. Leukemia 30, 1204–1208 (2016).

44. J. Chen et al., Cytokine receptor signaling is required for the survival of ALK-anaplastic large cell lymphoma, even in the presence of JAK1/STAT3 mutations. Proc Natl Acad Sci U S A 114, 3975–3980 (2017).

45. E. Masle-Farquhar et al., STAT3 gain-of-function mutations connect leukemia with autoimmune disease by pathological NKG2D(hi) CD8(+) T cell dysregulation and accumulation. Immunity 55, 2386–2404 e2388 (2022).

46. J. T. Warshauer et al., A human mutation in STAT3 promotes type 1 diabetes through a defect in CD8+ T cell tolerance. J Exp Med 218, (2021).

47. C. Gong et al., Sequential inverse dysregulation of the RNA helicases DDX3X and DDX3Y facilitates MYC-driven lymphomagenesis. Mol Cell 81, 4059–4075 e4011 (2021).

48. A. L. Lennox et al., Pathogenic DDX3X Mutations Impair RNA Metabolism and Neurogenesis during Fetal Cortical Development. Neuron 106, 404–420.e408 (2020).

49. T. Sengoku, O. Nureki, A. Nakamura, S. Kobayashi, S. Yokoyama, Structural basis for RNA unwinding by the DEAD-box protein Drosophila Vasa. Cell 125, 287–300 (2006).

50. H. Song, X. Ji, The mechanism of RNA duplex recognition and unwinding by DEAD-box helicase DDX3X. Nat Commun 10, 3085 (2019).

51. O. R. Bandapalli et al., The activating STAT5B N642H mutation is a common abnormality in pediatric T-cell acute lymphoblastic leukemia and confers a higher risk of relapse. Haematologica 99, e188–192 (2014).

52. L. Jiang et al., Exome sequencing identifies somatic mutations of DDX3X in natural killer/T-cell lymphoma. Nat Genet 47, 1061–1066 (2015).

53. C. Kucuk et al., Activating mutations of STAT5B and STAT3 in lymphomas derived from gammadelta-T or NK cells. Nat Commun 6, 6025 (2015).

54. H. L. Rajala et al., Discovery of somatic STAT5b mutations in large granular lymphocytic leukemia. Blood 121, 4541–4550 (2013).

55. A. Roberti et al., Type II enteropathy-associated T-cell lymphoma features a unique genomic profile with highly recurrent SETD2 alterations. Nat Commun 7, 12602 (2016).

56. E. D. de Araujo et al., Structural and functional consequences of the STAT5B(N642H) driver mutation. Nat Commun 10, 2517 (2019).

57. B. Maurer et al., High activation of STAT5A drives peripheral T-cell lymphoma and leukemia. Haematologica 105, 435–447 (2020).

58. H. T. T. Pham et al., STAT5BN642H is a driver mutation for T cell neoplasia. J Clin Invest 128, 387–401 (2018).

59. L. Yang, R. Rau, M. A. Goodell, DNMT3A in haematological malignancies. Nat Rev Cancer 15, 152–165 (2015).

60. M. Stoeckius et al., Simultaneous epitope and transcriptome measurement in single cells. Nat Methods 14, 865–868 (2017).

61. A. H. Jonsson et al., Granzyme K(+) CD8 T cells form a core population in inflamed human tissue. Sci Transl Med 14, eabo0686 (2022).

62. M. E. B. FitzPatrick et al., Human intestinal tissue-resident memory T cells comprise transcriptionally and functionally distinct subsets. Cell Rep 34, 108661 (2021).

63. C. Cellier et al., Safety and efficacy of AMG 714 in patients with type 2 refractory coeliac disease: a phase 2a, randomised, double-blind, placebo-controlled, parallel-group study. Lancet Gastroenterol Hepatol 4, 960–970 (2019).

64. J. K. Grewal, A. Kassardjian, G. A. Weiss, Successful novel use of tofacitinib for type II refractory coeliac disease. BMJ Case Rep 15, (2022).

65. N. Nandi et al., Normalization of duodenal mucosa after treatment with Janus kinase (JAK) inhibitor in refractory celiac disease type 2. Clin Res Hepatol Gastroenterol 46, 101960 (2022).

66. P. H. R. Green, S. Paski, C. W. Ko, A. Rubio-Tapia, AGA Clinical Practice Update on Management of Refractory Celiac Disease: Expert Review. Gastroenterology 163, 1461–1469 (2022).

67. W. Vader et al., The HLA-DQ2 gene dose effect in celiac disease is directly related to the magnitude and breadth of gluten-specific T cell responses. Proc Natl Acad Sci U S A 100, 12390–12395 (2003).

68. A. Al-Toma et al., Human leukocyte antigen-DQ2 homozygosity and the development of refractory celiac disease and enteropathy-associated T-cell lymphoma. Clin Gastroenterol Hepatol 4, 315–319 (2006).

69. R. D. Mendes et al., PTEN microdeletions in T-cell acute lymphoblastic leukemia are caused by illegitimate RAG-mediated recombination events. Blood 124, 567–578 (2014).

70. E. Papaemmanuil et al., RAG-mediated recombination is the predominant driver of oncogenic rearrangement in ETV6-RUNX1 acute lymphoblastic leukemia. Nat Genet 46, 116–125 (2014).

71. J. F. Ludvigsson et al., Diagnosis and management of adult coeliac disease: guidelines from the British Society of Gastroenterology. Gut 63, 1210–1228 (2014).

72. P. Blombery et al., Multiple BCL2 mutations cooccurring with Gly101Val emerge in chronic lymphocytic leukemia progression on venetoclax. Blood 135, 773–777 (2020).

73. G. L. Ryland et al., Novel genomic findings in multiple myeloma identified through routine diagnostic sequencing. J Clin Pathol 71, 895–899 (2018).

74. M. N. Marsh, P. T. Crowe, Morphology of the mucosal lesion in gluten sensitivity. Baillieres Clin Gastroenterol 9, 273–293 (1995).

75. D. W. Ruff, D. M. Dhingra, K. Thompson, J. A. Marin, A. T. Ooi, High-Throughput Multimodal Single-Cell Targeted DNA and Surface Protein Analysis Using the Mission Bio Tapestri Platform. Methods Mol Biol 2386, 171–188 (2022).

76. S. Picelli et al., Full-length RNA-seq from single cells using Smart-seq2. Nat Protoc 9, 171–181 (2014).

77. M. Singh et al., Lymphoma Driver Mutations in the Pathogenic Evolution of an Iconic Human Autoantibody. Cell 180, 878–894 e819 (2020).

78. T. Stuart et al., Comprehensive Integration of Single-Cell Data. Cell 177, 1888–1902 e1821 (2019).

79. H. Zhao et al., CrossMap: a versatile tool for coordinate conversion between genome assemblies. Bioinformatics 30, 1006–1007 (2014).

80. H. Li, A statistical framework for SNP calling, mutation discovery, association mapping and population genetical parameter estimation from sequencing data. Bioinformatics 27, 2987–2993 (2011).

81. A. McKenna et al., The Genome Analysis Toolkit: a MapReduce framework for analyzing next-generation DNA sequencing data. Genome Res 20, 1297–1303 (2010).

82. M. A. Field, Detecting pathogenic variants in autoimmune diseases using high-throughput sequencing. Immunol Cell Biol 99, 146–156 (2021).

83. H. Li, R. Durbin, Fast and accurate short read alignment with Burrows-Wheeler transform. Bioinformatics 25, 1754–1760 (2009).

84. W. McLaren et al., Deriving the consequences of genomic variants with the Ensembl API and SNP Effect Predictor. Bioinformatics 26, 2069–2070 (2010).

85. A. R. Hamzeh, T. D. Andrews, M. A. Field, Detecting Causal Variants in Mendelian Disorders Using Whole-Genome Sequencing. Methods Mol Biol 2243, 1–25 (2021).

86. A. T. Lun, D. J. McCarthy, J. C. Marioni, A step-by-step workflow for low-level analysis of single-cell RNA-seq data with Bioconductor. F1000Res 5, 2122 (2016).

87. G. Finak et al., MAST: a flexible statistical framework for assessing transcriptional changes and characterizing heterogeneity in single-cell RNA sequencing data. Genome Biol 16, 278 (2015).

88. G. Korotkevich et al., Fast gene set enrichment analysis. bioRxiv, 060012 (2021).

89. J. Li et al., KIR(+)CD8(+) T cells suppress pathogenic T cells and are active in autoimmune diseases and COVID-19. Science 376, eabi9591 (2022).

90. H. Li et al., The Sequence Alignment/Map format and SAMtools. Bioinformatics 25, 2078–2079 (2009).

91. T. D. Andrews, Y. Jeelall, D. Talaulikar, C. C. Goodnow, M. A. Field, DeepSNVMiner: a sequence analysis tool to detect emergent, rare mutations in subsets of cell populations. PeerJ 4, e2074 (2016).

92. S. Koren et al., Canu: scalable and accurate long-read assembly via adaptive k-mer weighting and repeat separation. Genome Res 27, 722–736 (2017).

93. R. Vaser, I. Sovic, N. Nagarajan, M. Sikic, Fast and accurate de novo genome assembly from long uncorrected reads. Genome Res 27, 737–746 (2017).

94. J. Ye, N. Ma, T. L. Madden, J. M. Ostell, IgBLAST: an immunoglobulin variable domain sequence analysis tool. Nucleic Acids Res 41, W34–40 (2013).

95. L. Tian et al., Comprehensive characterization of single-cell full-length isoforms in human and mouse with long-read sequencing. Genome Biol 22, 310 (2021).

96. J. A. Carter et al., Single T Cell Sequencing Demonstrates the Functional Role of alphabeta TCR Pairing in Cell Lineage and Antigen Specificity. Front Immunol 10, 1516 (2019).

97. A. A. Eltahla et al., Linking the T cell receptor to the single cell transcriptome in antigen-specific human T cells. Immunol Cell Biol 94, 604–611 (2016).

98. L. Song et al., TRUST4: immune repertoire reconstruction from bulk and single-cell RNA-seq data. Nat Methods 18, 627–630 (2021).

