## Supplementary figures and images for "Expanded T cell clones with lymphoma driver somatic mutations in refractory celiac disease"

### Data S1.pdf

RCD2-1:

IEL fraction:

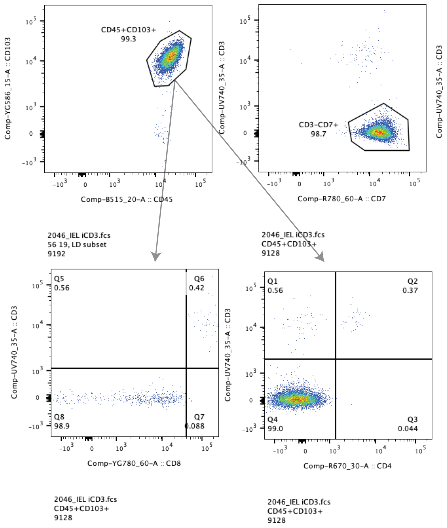


Lamina propria fraction:

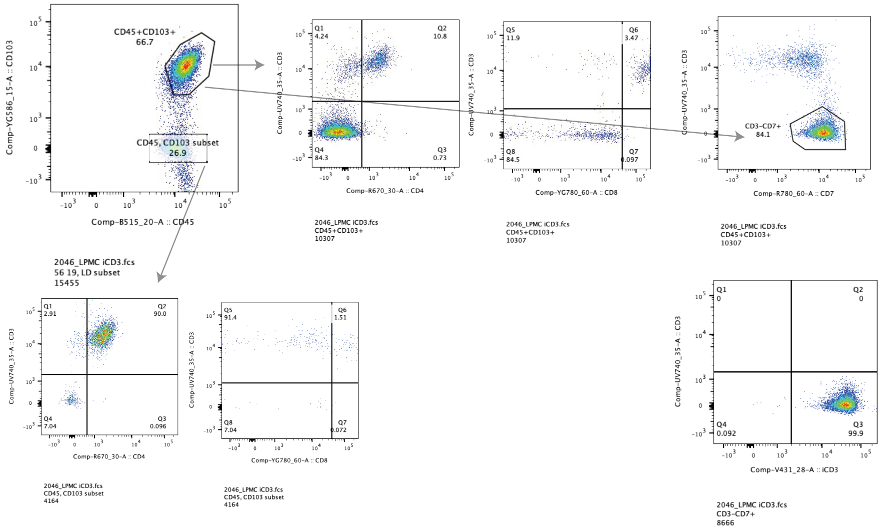


RCD2-2:

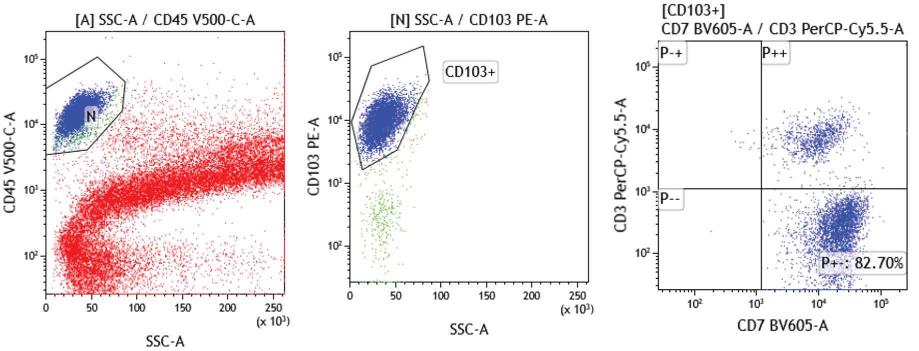


sCD3 stain

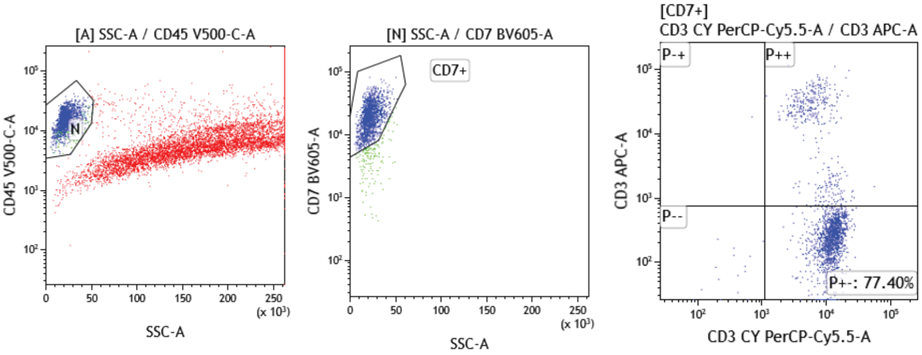


cCD3 stain

### Data S7.pdf

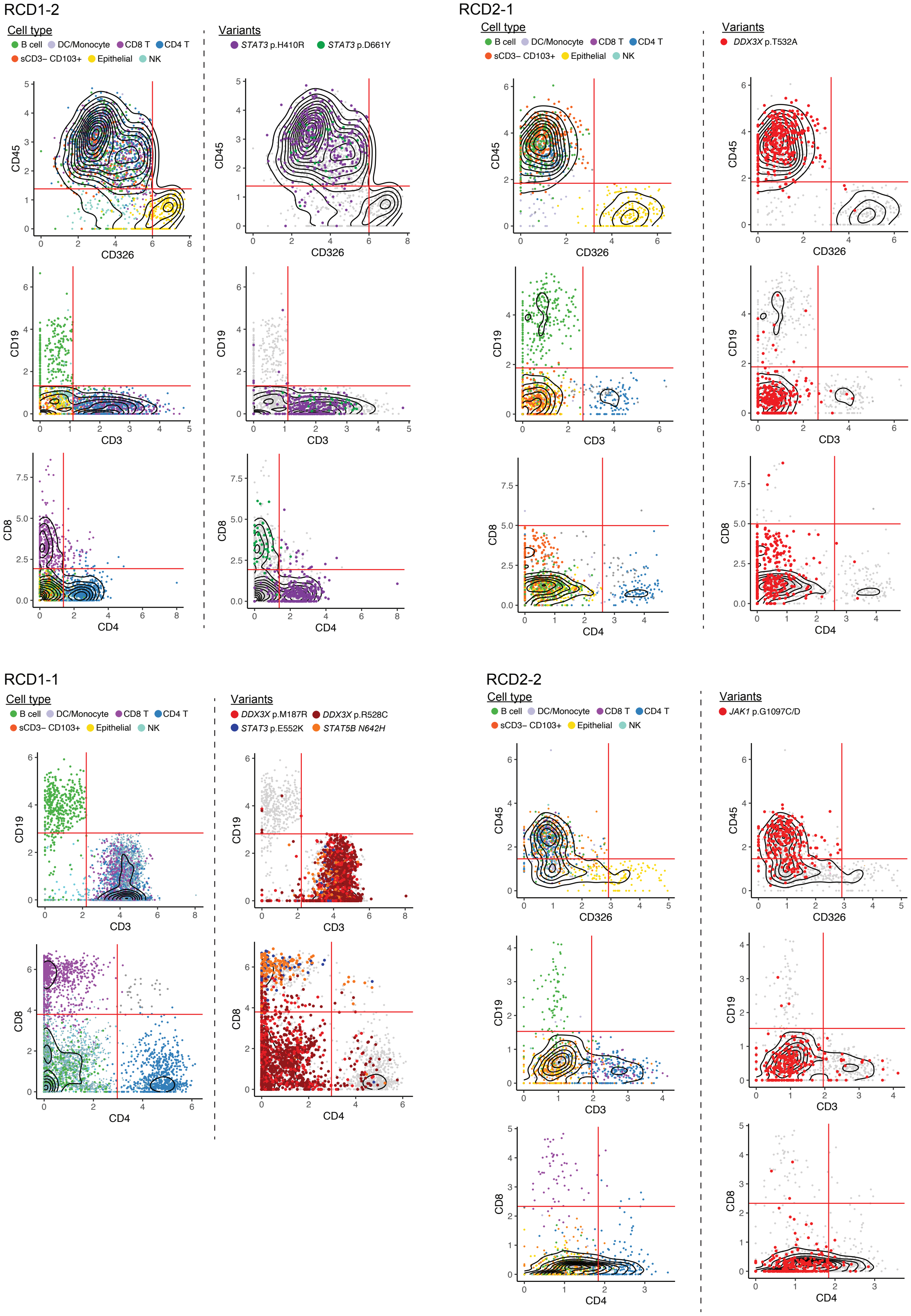
