## Supplementary materials for "Expanded T cell clones with lymphoma driver somatic mutations in refractory celiac disease"

Mandeep Singh *et al.*

**The PDF file includes:**

Materials and Methods
Supplementary text
Figs. S1 to S10
Tables S1 to S3

**Other Supplementary Materials for this manuscript include the following:**

Data S1 to S8

#### Materials and Methods

Human samples

According to internationally agreed criteria (*71*), diagnosis of CD was based on documented evidence of positive CD serology (anti-transglutaminase IgA antibody and anti-endomysial IgA antibody) in conjunction with villous atrophy and increased IELs (Marsh 3 enteropathy). RCD was defined by persistent villous atrophy (Marsh 3) after more than one year of strict adherence to a gluten-free diet (confirmed by expert dietitian review) and the presence of malabsorptive gastrointestinal symptomatology such as diarrhea and no other causes for symptoms or enteropathy found on clinical work-up. RCD2 was diagnosed based on evidence of an aberrant monoclonal IEL population using a combination of immunohistochemistry (>50% of IELs CD8-) and flow cytometry (>20% IELs CD45+ CD103+ sCD3- cCD3+ CD8-) (see table S1 and data S1). Targeted capture sequencing of total biopsy was also used to inform the diagnosis of RCD2-1, which was performed as previously described (*72, 73*) using the gene panel in data S5. RCD1 was diagnosed after excluding RCD2 i.e. normal IEL phenotype and the exclusion of any other enteropathies. See table S1 for clinical information for each individual and supplementary text for further clinical observations of RCD individuals RCD2-1, RCD2-2, and RCD1-1. Active newly diagnosed CD (ANCD) samples were collected from patients with positive CD serology at the time of their diagnostic endoscopy (with histology confirming CD) prior to them commencing a gluten-free diet. Healthy controls were individuals who had active CD excluded (negative CD serology and Marsh score <1).

Duodenal biopsies (typically, 6-8 from the 2nd part and 2 from the 1st part) were obtained during routine endoscopy at the Royal Melbourne Hospital, Melbourne Private Hospital (Melbourne, Australia), Blacktown Hospital (Sydney, Australia) or Fondazione IRCCS Policlinico San Matteo (Pavia, Italy). 4-6 2nd part duodenal biopsies were placed immediately in ice-cold RPMI, and then washed twice in PBS and placed in CryoStor CS10 freezing medium (Stemcell Technologies) and transported to liquid nitrogen cryogenic storage for long-term storage. For some individuals one 1st or 2nd part duodenal biopsy was collected for bulk DNA capture sequencing. The remaining duodenal biopsies were sent for routine histopathologic analysis and enteropathy grading where a Marsh score (*74*) was generated (Marsh 0 = normal, 1 = raised IELs, 2 = crypt hyperplasia, 3= villous atrophy with raised IELs and crypt hyperplasia). The study was conducted with approval from local Human Research Ethics Committees: Melbourne Health 2020.162 and Western Sydney Local Health District 2021/ETH01429.

Duodenal tissue biopsy digestion

Cryopreserved tissue biopsies were dissociated into single-cell suspensions by thawing the tissue in a 37 °C water bath followed by two washes with RPMI. The tissue was treated twice with 2mM EDTA with an incubation for 15min at 37 °C with continuous rotation. Supernatants containing the epithelial fractions were combined, washed, and kept on ice. The remaining tissue was treated with 1 U/mL Collagenase IV and 0.05 mg/mL of deoxyribonuclease (DNase) I in RPMI for 1h at 37°C with continuous rotation. Following digestion, remaining tissue was passed through a 18G needle multiple times and combined with the epithelial cell fraction. The combined mixture was filtered through a 100 μm cell strainer followed by cell counting and assessment of viability with Trypan Blue.

Flow cytometry sorting

Singe cell suspensions from duodenal biopsies were stained with antibodies against CD45, CD326 (EPCAM), CD3, CD4, CD8, CD19 and CD103. For G&T-Seq experiments, individual cells were sorted into 96-well LoBind PCR plates (Eppendorf) on a FACS Aria III (BD Biosciences). The following CD45+ cell types were sorted for G&T-Seq: RCD1-1, single CD3+ CD4- CD8- or CD3+ CD8+ cells; RCD1-2, single CD3+ CD8+ or CD3+ CD4+ cells; RCD1-6, single CD3+ CD8+ cells; RCD2-2, single CD3- CD19- cells. For single cell sequencing with the 10X Genomics platform, CD45+ were sorted in bulk and dead cells excluded based on viability dye staining (DAPI). Flow cytometry sorting was not performed for samples used with scDNA-Seq, except sample RCD1-1a where live CD45+ cells were sorted for.

Droplet-based cell surface protein and scDNA-Seq (MissionBio)

Single cell suspensions obtained from tissue biopsies were blocked with Human TruStain FcX (BioLegend) for 15 min on ice and then incubated with Total-Seq D antibodies (Biolegend) for 30 min. See data S6 (sheet: "Total-Seq D panel – scDNA-Seq") for a list of TotalSeq-D antibodies used. After three washes with cell staining buffer (Biolegend), cells were filtered through a 40 μm strainer, counted with a hemacytometer, and then loaded into the Tapestri instrument (MissionBio) according to the manufacturer's protocol (*75*). Briefly, single cells were encapsulated into droplets containing a lysis buffer and incubated for 1 h at 50 °C followed by 10 min at 80 °C. Droplets containing single-cell barcoding beads were combined with the encapsulated cell lysates and a panel of primers designed to amplify either 12 driver genes (exonic regions) comprising of 269 amplicons (panel 1), or 54 driver genes (exonic regions and hotspots) comprising of 557 amplicons (panel 2). The designed amplicons covered 81.94% of the target regions in panel 1 and 89.7% of the target regions in panel 2. The droplets were exposed to ultraviolet light to cleave the PCR primers containing unique cell barcodes from the beads and PCR was performed for 23 cycles with temperature gradients recommended by the manufacturer. Amplified products were purified using AMPure XP beads (Beckman Coulter) and the protein tag library separated using magnetic isolation using streptavidin beads and a biotinylated oligonucleotide complementary to the 5' end of the antibody tags. Additional PCR was carried out using Illumina index primers on DNA and protein libraries. The libraries were sequenced on an Illumina NovaSeq instrument with 150bp paired end-reads to the depth recommended by the manufacturer. Data S2 (sheets: "DNA panel 1", "DNA panel 2" and "Sample scDNA-Seq details") details the panel design, target regions, number of cells loaded for each sample, and sequencing coverage.

Plate based single cell gDNA and mRNA sequencing (G&T-Seq)

gDNA and mRNA were isolated from single cells sorted into 96-well plates using the G&T-Seq protocol (*37*). The mRNA of each cell was processed for scRNA-Seq using the Smart-seq2 protocol (*76*), with modifications described previously (*77*). Smart-seq2 mRNA sequencing libraries were sequenced using an Illumina NextSeq 500 instrument with 150 bp paired-end reads to a median depth of ~1 million reads per cell. gDNA was subjected to multiple displacement amplification using the REPLI-g Single Cell Kit (QIAGEN) according to the manufacturer's instructions and purified using AMPure XP beads (Agencourt). Gene-specific PCR amplification was performed by diluting gDNA 1/100 and using primers outlined in data S6 (sheet: "Primers") and the Taq polymerase kit (Invitrogen) with the following conditions: 95°C for 3 min; [95°C for 30 s, 60°C for 15 s, 72°C for 1 min] x 35 cycles; 72 °C for 5 min. Positive PCR products were Sanger sequenced by the Garvan Molecular Genetics facility (Garvan Institute, Australia).

Droplet-based cell surface protein, TCR and scRNA-Seq (10X Genomics)

Flow sorted CD45+ cells from tissue biospies were blocked with Human TruStain FcX (BioLegend) for 15 min on ice and then incubated with 159 Total-Seq A antibodies (Biolegend) for 30 min on ice. See data S6 (sheet: "Total-Seq A panel – scRNA-Seq") for the list of TotalSeq-A antibodies used. After three washes with PBS + 2% FCS, cells were filtered through a 40 μm strainer and counted with a hemacytometer. scRNA-Seq libraries were prepared using the Chromium Single Cell 3' v3 protocol (10X Genomics) according to the manufacturer's protocol, aiming for a recovery of 10,000 - 15,000 cells for each sample. Briefly, single cells were encapsulated into droplets in the Chromium Controller instrument for cell lysis and barcoded reverse transcription of mRNA and tagging of oligo barcoded antibodies, followed by amplification, shearing and Illumina library construction. An Illumina NovaSeq instrument (150 bps, paired-end reads) was used to sequence the libraries at a depth of ~50,000 reads per cell for scRNA-Seq libraries and ~30,000 reads for protein-tag libraries. Full-length TCRα, TCRβ, TCRδ and TCRγ sequences were obtained from the scRNA-Seq library using RAGE-Seq (*36*). Between 50-100 ng of full-length cDNA was used for targeted enrichment of all functional TCR and BCR genes with the xGen NGS Hybridization Capture protocol (Integrated DNA Technologies), according to the manufacturer's recommendations. The following modifications were included: xGen blocking oligos were not used, hybridisation time was increased to 16 h, and PCR was used with primers that amplify full-length cDNA (Forward: AAGCAGTGGTATCAACGCAGAGT; Reverse: CTACACGACGCTCTTCCGATCT) and the PCR extension time increased to 90 s.

Functional assays of STAT3 variants

For protein activation assays, the A4 $STAT3^{-/-}$ colon cancer cell line, kindly provided by Jean-Laurent Casanova, was seeded at 500,000 cells per well and grown for 24 h before being transfected with either an expression vector containing human *STAT3* cDNA (NM_139276), variant *STAT3* or empty vector using the Lipofectamine 3000 kit (Thermofisher) akin to manufacturer's instructions. All expression vectors shared the DDK-Myc-pCMV6 backbone (Origene, #RC215836) with mutations introduced in-house using site-directed mutagenesis. After 24 h of transfection, cells were either mock stimulated (serum-free McCoy media) or stimulated with 100 ng/mL IL-6/IL-6Rα chimera (R&D) for 15 min before being washed with serum-free McCoy media and rested for 1 or 2 h to assess de-phosphorylation of STAT3. Cells were scraped and prepared in cell lysis buffer (150 mM NaCl, 50 mM Tris, pH 8, 5 mM EDTA, 1% Triton, 10 mM DTT) supplemented with a protease inhibitor cocktail (Roche) for 15 min on ice followed by centrifugation at 13,000 x g for 10 min to remove debris. Protein extracts were analyzed via western blot using the Bio-Rad system. Briefly, 50 ug of protein extract was calculated using a Bradford Assay (Bio-Rad) and run on an 7.5% polyacrylamide gel before being transferred to a PVDF membrane and blotted for total STAT3 (Cell Signaling Technology), Tyr705-phosphorylated STAT3 (Cell Signaling Technology) and loading control GAPDH (Santa-Cruz). Membranes were fluorescently imaged using the LI-COR Odyssey CLx and analyzed by ImageStudio (LI-COR Biosciences).

Transcriptional activity of variants was assessed using the Dual-Luciferase Reporter Assay System (Promega) following the manufacturer's instructions. A4 $STAT3^{-/-}$ cells were co-transfected with a reporter vector (pGL 4.47) containing five-copies of the luciferase reporter gene, *Luc2P*, downstream of the sis-inducible element and a constitutively expressed vector containing *Renilla* luciferase (pGL 4.74). Expression vectors containing wildtype STAT3, variant STAT3 or empty expression vector were also co-transfected and incubated for 24 h before being mock stimulated or stimulated with 50 ng/mL IL-6/IL-6Rα for a further 24 h. Cells were then lysed before firefly and renilla luciferase substate was sequentially introduced and luminescence read on a CLARIOstar plate reader. The ratio of firefly to renilla luminescence was used to calculate relative luciferase activity due to STAT3.

scDNA-Seq surface protein data analysis

FASTQ files from the scDNA-Seq protein libraries were run through the Tapestri Pipeline (v2, Mission Bio), to align reads to the antibody barcode list, and assign antibody reads to cell barcodes identified from the DNA data detailed below. For each sample, the extracted protein counts matrix was normalised in Seurat v4 (*78*) using a centred log ratio transformation (CLR) with the *NormalizeData(normalization.method="CLR", margin=2)* function. Following this, PCA (using *RunPCA()*) dimensionality reduction and neighbour (using *FindNeighbours()*) calculation were performed on each sample independently, and unsupervised clusters (using resolutions ranging from 0.5 to 1.2) were identified using the *FindClusters()* function. Cell type annotation was then performed manually based on the protein expression in each cluster using the canonical markers outlined in data S2 (sheet: "cell type classification"). Cell types that were determined to be "dead", "doublet" or "unknown" were removed. A custom supervised cell type annotation algorithm was developed to optimise protein marker thresholds for six key markers: CD45, CD3, CD4, CD8, CD19, CD326. This supervised annotation method was used to further refine cell types by further filtering out dead, doublet and unknown cells. See fig. S1A for an outline of this process, and fig. S1B to D which describes the number of cells filtered at each step and the proportions of cell types before and after filtering.

Cells identified as CD45+ CD19- CD3- (found in 10/23 individuals) were further classified as either "sCD3- CD103+ cells" or "sCD3- CD103- cells" based on high or low expression of CD103 and CD7 (*14*), respectively (fig S1F). For experiments involving samples RCD2-1 and RCD1-2, the CD103 Total-Seq D antibody was not included so high expression of CD7 alone was used to identify the aberrant sCD3- CD103+ cells. The identity of sCD3- CD103- cells likely consist of misassigned cells and plasmablasts with low to negative CD19 expression since the UMAP in fig. S1G shows the majority of these cells sit in the B cell cluster.

The integrated UMAP of all 23 samples (Fig. 1) was generated using the suggested integration pipeline from Seurat (*78*). The *SelectIntegrationFeatures()* function was used to select features that were repeaded variable across samples, and then used to find anchors using *FindIntegrationAnchors()*. The data was then integrated using this anchor set with the function *IntegrateData(dims = 1:20*). Dimensionality reduction was then performed to produce a PCA (using RunPCA()) and subsequent UMAP (using *RunUMAP(reduction="pca", min.dist=0)*).

scDNA-Seq variant data analysis

FASTQ files from the scDNA-Seq DNA libraries were analysed with the Tapestri Pipeline (v2, Mission Bio), to align reads to the human genome (GrCh37), perform barcode correction and assign sequence reads to cell barcodes. The alignment files generated from Tapestri Pipeline were converted from GRCh37 assembly to GRCh38 using CrossMap (*79*). Sequencing coverage for each amplicon was determine using the SAMtools (*80*) depth command for all cells and GATK (*81*) DepthofCoverage for cell type specific calculations (detailed in fig. S2A to B).

Variants were called using a modified version of previously described variant detection workflow (*82*) which utilizes BWA (*83*) for alignment, GATK best practices for group variant calling (*81*) and variant effect predictor for annotation (*84*). Variants were annotated as previously described (*85*) along with cell annotations obtained from protein markers (as described above). Allele frequencies of each variant for each cell type were calculated and compared to the total allele frequency. Variants were retained if they met the following criteria: the variant constitutes either a nonsynonymous, nonsense or frameshift mutation, it is present in more than 5 total cells, it contains an average variant quality score greater than 100, it has genotyping calls in more than 50% of cells, it has a mean allele frequency greater than 0.4. Following this initial filtering, approximately 20-50 variants remained per sample. To further identify true positive somatic variants, these remaining variants were interrogated within each cell type where variants were required to belong to more than 80% of cells of a specific cell type amongst the major cell populations: CD4, CD8, B cells, and Epithelial cells (and CD3- CD103+ cells or CD3+ CD4- CD8- cells if present). To account for variants that may arise in hematopoietic progenitor cells, variants had to belong to more than 20 cells in total and present in less than 10% of epithelial cells. fig. S2D demonstrates the identification of the final variants using this approach for each individual sample where somatic mutations were found. A list of final variants is shown in table S2. Allele dropout was calculated by identifying germline heterozygous variants that were present in more than 500 cells and were assigned a reference single nucleotide polymorphism (rs) number in dbSNP, and had genotyping calls in more than 90% of cells. The number of wild-type and homozygous calls for that variant were then calculated as a fraction of total genotype calls (fig. S2C).

To further interrogate cell type assignment for those samples with somatic mutations, major protein markers were plotted against each other (CD45 vs EPCAM, CD19 vs CD3, CD8 vs CD3) for each mutant clone and compared to wild-type cells. Mutant cells that were assigned an alternative cell type were often found to contain low marker expression of the dominant cell type. This suggests that the misassigned mutant cells are likely due to technical reasons. This is demonstrated in data S7 for samples RCD2-1, RCD2-2, RCD1-1 and RCD1-2. The percentage of misassigned variant cells is also in concordance with the expected doublet rate of ~5%.

<u>scRNA-Seq gene expression and surface protein data analysis</u>

Gene expression matrices corresponding to unique molecular identifier (UMI) counts from 10X scRNA-Seq libraries were produced using the Cell Ranger workflow (v3.0.1, 10X Genomics), which included alignment to the GRCh38 reference genome, quality control and UMI counting. Paired protein matrices were produced using the CITE-seq-Count tool (https://github.com/Hoohm/CITE-seq-Count). Samples underwent quality control to remove cells with low mitochondrial content, and low/high number of unique genes and total counts. Gene expression was normalized using Scran (*86*) with default parameters, while protein expression was normalised using centred-log-ratio.

Gene and protein data for each sample were integrated together using the weighted nearest neighbour method implemented in the Seurat package v4 (*78*). First, the 2000 most variable genes were calculated using *FindVariableFeatures*, followed by scaling using the *ScaleData* function and PCA using *RunPCA*. *FindMultiModalNeigbours* was then used to find the cell nearest neighbour graph based on both the gene and protein data. This graph was then used for subsequent clustering using *FindClusters* and UMAP generation using *RunUMAP* using nearest neighbour=30 and min_dist=0.01. Longitudinal samples for RCD2-1 (2 timepoints) and RCD1-1 (3 timepoints) were integrated for each sample to control for batch effects using the recommended pipeline in the Seurat v4 package (*78*). Gene expression matrices were aggregated using the *FindIntegrationAnchors* and *IntegrateData* functions. The resulting integrated matrix was then utilised to perform dimensionality reduction and clustering. First, the top 3000 highly variable genes were extracted using the *FindVariableFeatures* function. Principal component analysis was performed using the *RunPCA* function with the option npcs=30 followed by neighbour and cluster calculation using *FindNeighbours* and *FindClusters* functions respectively. Based on the integrated data, UMAP generation was obtained by *RunUMAP* using nearest neighbour=30 and min_dist=0.01. Cell annotation was performed manually by interrogating resolution parameters from 0.01 to 1.5 to determine the marker genes and proteins for each resolution. The resolution parameter was then chosen after manual inspection of the marker genes at each resolution level. For RCD1 samples RCD1-1, RCD1-2 and RCD1-6, cell clusters that were not positive for protein CD3 were not plotted on the final UMAPs. This was also performed for the two RCD2 samples RCD2-1 and RCD2-2, with the exception that sCD3- CD103+ cells were retained. The list of cell clusters retained and removed from the final UMAP analysis is detailed in data S3 (sheet: "scRNA-Seq – cell types"). Excluded cell clusters mainly comprised of B cells and plasmablasts.

Differential expression analysis for genes and proteins

Differential gene and protein expression analysis of scRNA-Seq data was performed using the *FindMarkers* function in Seurat v4 (*78*) using the method MAST (*87*), which uses a hurdle model to identify statistically significant genes based on the number and expression levels of genes between two groups of cells. For differential protein expression, the *FindMarkers* function was used using the Wilcoxon Sum Rank test. Pairwise cluster differential analysis was performed for both gene and protein data, with cluster markers determined using the median p-value and median fold change formed from the pairwise analysis. Only significant genes and proteins were used to determine the median p-values and fold changes, with adjusted p-value < 0.01 and log2FC > 0.014. Dot plots to visualise the results of differential expression were generated using ggplot2, and volcano plots generated using the *EnhancedVolcano* function in the EnhancedVolcano R package. The full list of differentially expressed genes and proteins from the pairwise analysis used to generate volcano plots are detailed in data S3. Gene set enrichment analysis (GSEA) was performed using the *fgsea* function in the fgsea R package (*88*) with default parameters, based on DGE results ranked by the fold-change. Gene sets are detailed in data S6 (sheet: "GSEA gene lists"). Gene sets include differentially expressed genes between CD8 GZMK+ and GZMB+ cells ("Jonnson et al. GZMK+" and "Jonnson et al. GZMB") described in inflamed human tissue (*61*), differentially expressed genes between CD8 CD103+ and CD8 CD103- cells ("FitzPatrick" et al. CD103+" and "FitzPatrick et al. CD103-") described in healthy human intestine (*62*), and genes enriched in CD8 KIR+ cells compared to CD8 KIR- cells ("CD8: KIR+ vs KIR-") described by Li et al. (*89*).

Analysis of *JAK1* LOH from scRNA-Seq data

For each cell type annotation, the original BAM file was subsetted using 10X Genomics subset-bam software (https://github.com/10XGenomics/subset-bam). Samtools pileup (*90*) was used to extract raw sequence data for both the *JAK1* and a comparably sized *STAT3*/*STAT5b* regions with the latter used as a control. For each cell, custom code previously described (*91*) was used to extract all coordinates with at least two variant reads and variant:reference ratios calculated to determine whether variants were likely homozygous or heterozygous. Details of the larger workflow are available at https://github.com/acalcino/cnv_caller.

<u>TCR reconstruction and analysis</u>

For samples RCD1-1 (all timepoints), RCD1-2, and RCD1-6, processing of RAGE-Seq data was performed as previously described (*36*). Briefly, DNA amplicons were sequenced using Oxford Nanopore Technologies' (ONT's) ligation sequencing kit SQK-LSK109 on ONT's R9.4.1 flowcells (FLO-PRO002). Basecalling was performed with ONT's Guppy software using major version 4. The base-called FASTQ files were de-multiplexed using the 16 nucleotide (nt) 10X cell barcodes by scanning the first and last 200 nt of any read longer than 250 nt for a matching sequence, with <2 mismatches. All reads assigned to a given cell were then assembled *de novo* using Canu (v1.8) (*92*) and sequence-polished using Racon (v1.3.3) (*93*). The polished contigs were analyzed with IgBlast (*94*) to determine V(D)J sequences of TCRα, β, δ and γ chains. T cell clones were defined as cells carrying the same paired TCRα and TCRβ V, J and CDR3 sequence. For samples RCD2-1 and RCD2-2, RAGE-Seq data was run through a pipeline that extracts 10X UMI sequences. The match_cell_barcode tool within FLAMES v0.1 (*95*) was used to demultiplex base-called FASTQ files and to extract the barcode and candidate UMI sequence for each read. UMI sequences were then clustered to the most common UMI for each barcode with a maximum edit (Levenshtein) distance of one. UMIs represented by less than 5 constituent reads were excluded. The constituent reads for each UMI were assembled *de novo* using the RAGE-Seq pipeline above. TCRγ chains were required to have a UMI count >2. The RAGE-Seq output is shown in data S8. The healthy control CD4 and CD8 dataset used for comparison against the RCD2 clones in fig. S4D-G is sourced from Carter et al. (96), with the mean calculated across donors.

TCRα, β, δ and γ chains were assembled from plate-based Smart-Seq2 sequencing data from G&T-Seq with VDJPuzzle (*97*) or TRUST4 (*98*) and are detailed in data S4.

**Supplementary Text**

Below are clinical notes on the diagnosis of individuals RCD2-1, RCD2-2, and RCD1-1.

RCD2-1

CD3+ IEL have >50% loss of CD8 expression; CD4+ increased on lamina propria. Individual presented with severe diarrhea and weight loss; improved with immunosuppression and cladribine.

RCD2-2

Individual RCD2-2 displayed an increase in IELs, present in all sites but more significant in the fragments of duodenum-bulb and II duodenal portion. IELs were small in size and otherwise atypical, characterized by an altered phenotype with positivity for CD3 and CD7 and negativity for CD4, CD5, CD8, not expression of CD30 and CD56. Staining for TCRAB and TCRGD showed negativity intraepithelially for TCRAB and rare TCRGD positive elements (lower than CD3+ cells). The component of the lamina propria showed no abnormal phenotype. EBV was negative (contrary to the original laboratory result, in which probably cytoplasmic positivity to in situ hybridization was documented). The observation has not allowed at the time to define the clearly lymphomatous lesion but were consistent with a picture of refractory celiac disease type 2 (ulcerative jejunitis). TCR gamma rearrangement was polyclonal.

RCD1-1

The diagnosis of individual RCD1-1 was the subject of considerable discussion between gastroenterology and hematology specialists. The individual had persistent enteropathy despite a prolonged, strict gluten-free diet but was minimally symptomatic, which is atypical for RCD. Duodenal flow cytometry showed an atypical infiltrate of small lymphocytes in the lamina propria, with 54% being CD2+, sCD3+, CD4-, CD5+, CD7+/- (variable), CD8-, CD10- and CD16/56-; however no aberrant IEL sCD3- CD103+ clones were detected, meaning they were not classified as RCD2. However, the combination of persistent enteropathy non-responsive to a trial of steroid therapy and the T cell infiltrate led to the treatment with the purine analogue chemotherapeutic agent cladribine, traditionally reserved for RCD2 patients and he has remained clinically stable since.

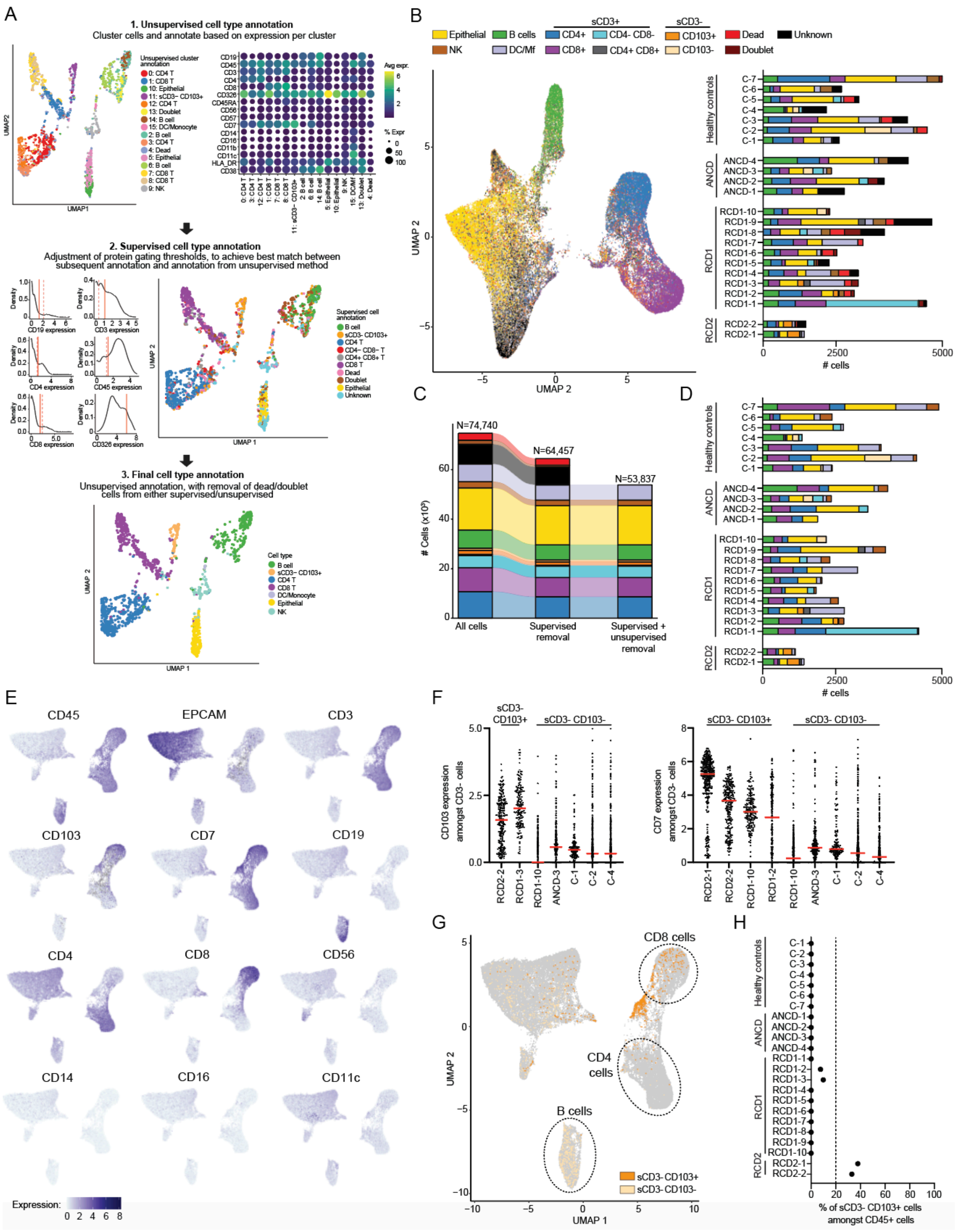


**Fig. S1. Cell type identification from scDNA-Seq cell surface protein data**

(**A**) Schematic of the cell type annotation process of scDNA-Seq protein data to identify cell types (shown for individual RCD2-2). In step 1, unsupervised cell type annotation involves dimensionality reduction (UMAP) and clustering of cells based on their normalized protein expression values (top left panel). The cell type of each cluster is annotated manually based on the expression of key markers (top right panel). Cell populations identified as “dead”, “doublet” and “unknown” are removed. In step 2, supervised cell type annotation involves manually selecting initial values for gating key protein markers (middle left panel, with dashed red lines representing initial gating values), and subsequent cell type calling based off these thresholds (middle right panel). The following six protein markers were used for the supervised analysis: CD19, CD8, CD4, CD3, CD45 and CD326. Gating thresholds are then adjusted (solid red lines in middle left panel) to maximize congruence with the unsupervised cell annotation. In step 3, doublet, dead and unknown cells identified by the supervised annotation are further filtered out from the unsupervised annotation (bottom panel). (**B**) Left panel, UMAP of duodenal cells of 23 individuals integrated together using 83 cell surface proteins prior to the removal of “dead”, “doublet” and “unknown” cell types by unsupervised and supervised analysis. Right panel, number of cells belonging to different cell types for each sample. n=74,740 total cells. (**C**) Alluvial plot showing the number of cells belonging to each cell type amongst all samples (n=23) at each step of removal of “dead”, “doublet” and “unknown” cell types using supervised and unsupervised analysis. Colors correspond to cell types shown in (B). The total number of cells across all samples is shown above. (**D**) Number of cells belonging to each cell type for each individual sample following the removal of “dead”, “doublet” and “unknown” cell types by both unsupervised and supervised methods. n= 47,946 total cells. (**E**) Imputed cell surface protein expression on the final integrated UMAP (Fig. 1A) for selected markers that were used for cell type annotation. (**F**) Expression (normalized reads) of surface CD103 (left panel) and CD7 (right panel) of cells classified as CD45+ CD19- CD3- in the depicted individuals. Red lines show the median. Number of CD45+ CD19- CD3- cells: RCD2-1, 370; RCD2-2, 234; RCD1-2, 210; RCD1-3, 166; RCD1-10, 361; ANCD-3, 190; C-1, 142; C-2, 381; C-4, 381. CD19- CD3- cells are further classified as sCD3- CD103+ or sCD3- CD103- based on CD7 and CD103 expression which are indicated above each sample. (**G**) UMAP as in Fig. 1A highlighting only the sCD3- CD103- and sCD3- sCD103+ cell types. The CD8, CD4 and B cell clusters are highlighted with dashed circles. (**H**) Proportion of sCD3- CD103+ cells amongst total CD45+ cells for each individual sample. sCD3- CD103+ cells are only detectable in samples RCD2-1, RCD2-2, RCD1-2 and RCD1-3. Dashed vertical line indicates the 20% cut-off of sCD3- CD103+ cells required for RCD2 diagnosis.

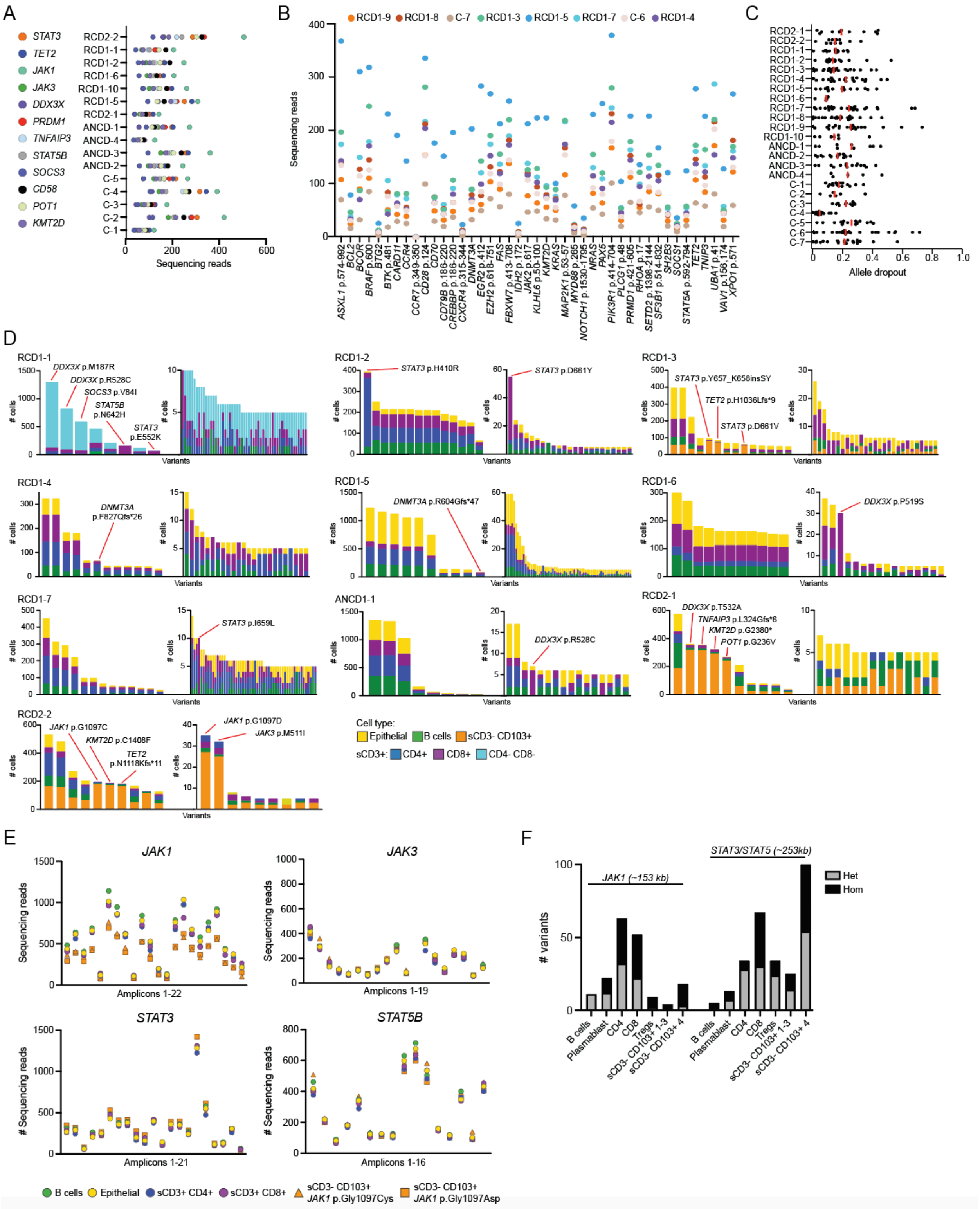


**Fig. S2. Variant analysis of scDNA-Seq data**

(**A-B**) Sequencing coverage of the genes (exonic regions) and selected gene regions (mutation hotspots) used in the scDNA-Seq amplicon panels for each individual sample. Genes common to both panels 1 and 2 are shown in (A), while genes and gene regions present only in panel 2 are in shown in (B). Panel 1 mean coverage (± sdev) for each disease group: RCD2, 261 ± 106; RCD1, 127 ± 55; ANCD, 141 ± 72; healthy controls (C), 131 ± 88. Panel 2 mean coverage (± sdev): RCD1 (n=6), 111 ± 85; controls, 60 ± 44. (**C**) Allele dropout of germline heterozygous variants present in >500 cells for each individual sample. Allele dropout was calculated as the fraction of wild-type and homozygous calls amongst total genotype calls. Red lines show the median. Median allele dropout (± sdev) for each disease group: RCD2, 0.17 ± 0.13; RCD1, 0.17 ± 0.14; ANCD, 0.21 ± 0.12; healthy controls (C), 0.20 ± 0.14. A total of 347 germline variants were queried across the 23 samples. (**D**) Cell type proportions for each variant following initial variant filtering shown for individual samples where somatic mutations were identified. Final filtered variants for each sample are indicated with red lines along with the name of the variant. The final variants were required to be present in more than 5 cells and belonging to >80% of a specific cell type or present in more than 20 cells across multiple immune cell types but consisted of <10% epithelial cells. The cell types CD4, CD8, B cells and epithelial cells (and sCD3- CD103+ or sCD3+ CD3- CD8- cells if present) were used for this analysis. Number of variants following initial filtering: RCD1-1, 61; RCD1-2, 37; RCD1-3, 42; RCD1-4, 36; RCD1-5, 75; RCD1-6, 27; ANCD1-1, 24; RCD2-1, 24; RCD2-2, 19. Bar plots are split across two plots for visualization purposes. (**E**) Mean sequencing coverage of each amplicon of *JAK1*, *JAK3*, *STAT3*, *STAT5B* genes for the indicated cell types for individual RCD2-2. sCD3- CD103+ cells are further split into the two clones with different driver mutations (*JAK1* 1097C vs *JAK1* 1097D). The number of amplicons per gene is indicated on the x-axis. Amplicons were required to have a mean coverage of >50 sequencing reads across all cell types. Number of cells: B cells, 104 cells; Epithelial, 131 cells; sCD3+ CD4+, 203 cells; sCD3+ CD8+, 60 cells; sCD3- CD103+ *JAK1* 1097C, 146 cells; sCD3- CD103+ *JAK1* 1097D, 15 cells. *JAK1* median coverage (± sdev): B cells, 603 ± 300; Epithelial, 574 ± 284; sCD3+ CD4+, 516 ± 275; sCD3+ CD8+, 540 ± 258; sCD3- CD103+ *JAK1* 1097C, 378 ± 185; sCD3- CD103+ *JAK1* 1097D, 394 ± 190. *JAK3* median coverage (± sdev): B cells, 157 ± 186; Epithelial, 132 ± 169; sCD3+ CD4+, 130 ± 168; sCD3+ CD8+, 168 ± 183; sCD3- CD103+ *JAK1* 1097C, 149 ± 183; sCD3- CD103+ *JAK1* 1097D, 196 ± 181. *STAT3* median coverage (± sdev): B cells, 303 ± 262; Epithelial, 277 ± 257; sCD3+ CD4+, 244 ± 247; sCD3+ CD8+, 280 ± 263; sCD3- CD103+ *JAK1* 1097C, 278 ± 250; sCD3- CD103+ *JAK1* 1097D, 300 ± 285. *STAT3* median coverage (± sdev): B cells, 201 ± 204; Epithelial, 200 ± 196; sCD3+ CD4+, 194 ± 180; sCD3+ CD8+, 192 ± 191; sCD3- CD103+ *JAK1* 1097C, 189 ± 175; sCD3- CD103+ *JAK1* 1097D, 206 ± 203. (**F**) Proportion of heterozygous (Het) and homozygous (Hom) variants called from scRNA-Seq 10X data of individual RCD2-2 (see Fig. 2) within genomic regions covering *JAK1* (chr1: 64823229-64976549) or *STAT3* and *STAT5* (chr1: 64823229-64976549) gene loci. Cell types correspond to those annotated from the scRNA-Seq analysis. sCD3- CD103+ 1-3 clusters correspond to cells carrying *JAK1* 1097C and sCD3- CD103+ 4 cluster corresponds to cells carrying *JAK1* 1097D (see Fig. 2). Number of cells: B cells, 50; plasmablasts, 325; CD4, 2209; CD8 (individual clusters combined), 676; Tregs, 139; sCD3- CD103+ 1-3, 421; sCD3- CD103+ 83.

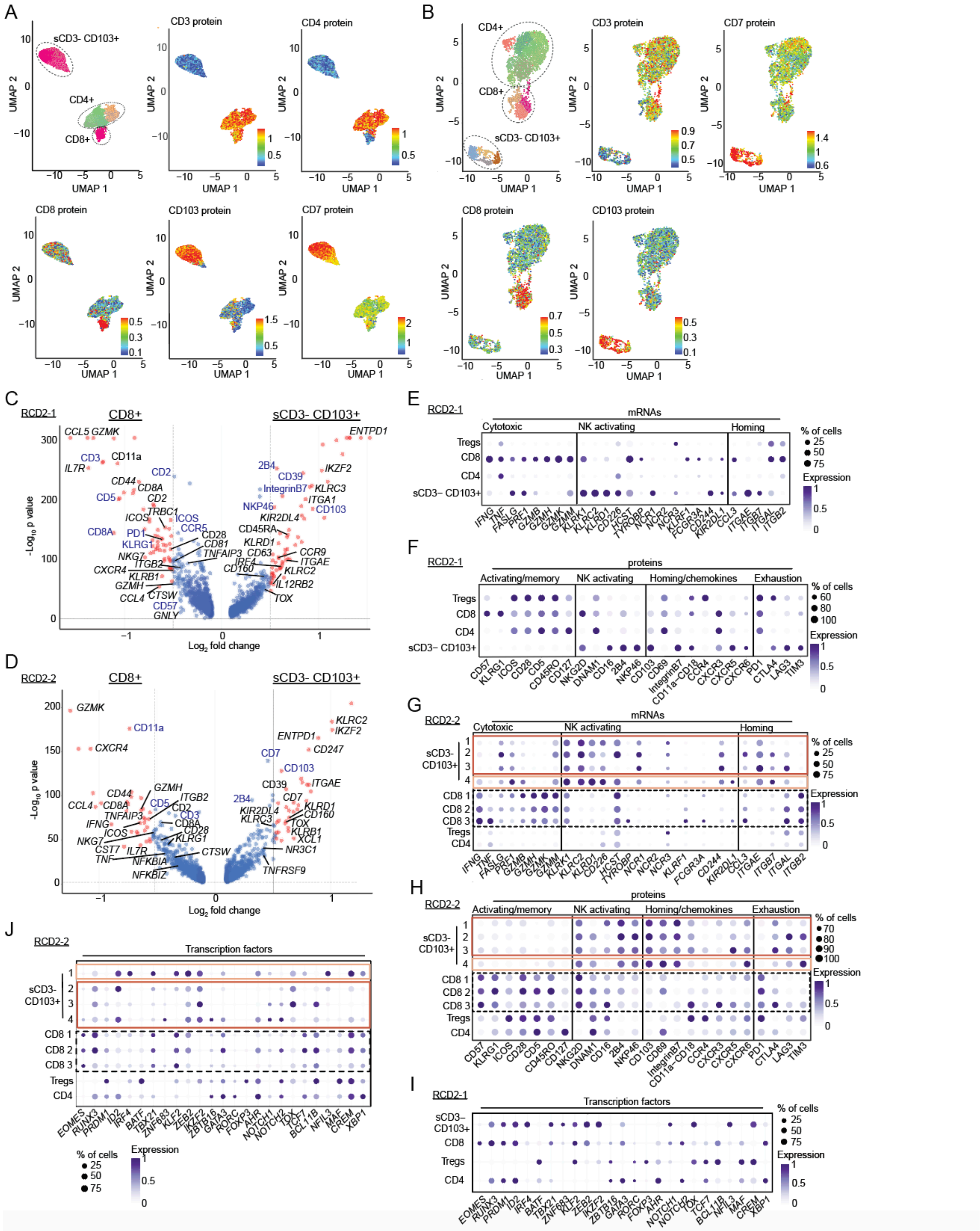


**Fig. S3. Cytotoxic and inflammatory gene programs of RCD2 sCD3- CD103+ cells**
(**A-B**) UMAP analysis of CD45+ sCD3+ and CD45+ sCD3- CD103+ cells for individual RCD2-1 (A) and individual RCD2-2 (B) using combined gene-expression and cell surface protein data. The major cell types sCD3- CD103+, CD4+ and CD8+ are indicated in the top left UMAP, while

the remaining UMAPs show the cell surface protein expression of selected markers. RCD2-1, n=4,661 cells (1760 CD4+ cells, 503 CD8+ cells, 2398 sCD3- CD103+ cells); RCD2-2, n=3,389 cells (2,209 CD4+ cells, 676 CD8+ cells, 504 sCD3- CD103+ cells). (**C-D**) Volcano plots of differentially expressed genes and proteins between CD8+ (n=503) and sCD3- CD103+ (n=2,398) cells for individual RCD2-1 (C), and between CD8+ (n=676) and sCD3- CD103+ (n=504) cells for individual RCD2-2 (D). Vertical dotted lines represent $\log_2$ fold change > 0.1. Proteins or genes that are significant are denoted in red and have a $\log_2$ fold change > 0.1 and p-value < 0.05. Differentially expressed gene names are annotated in black and differentially expressed protein names are annotated in blue. The full list of differentially expressed proteins and genes is described in data S3. (**E-J**) Dot plots of selected genes (E, G, I to J) and proteins (F, H) of the indicated cell type for individuals RCD2-1 (E to F, I) and RCD2-2 (G to H, J). Colored boxes mark sCD3- CD103+ cell clusters that carry clonal TCRγ chains depicted in Fig. 2C (RCD2-2). The size of each dot represents the percentage of cells with non-zero expression. The color intensity represents the scaled log expression averaged over all cells.

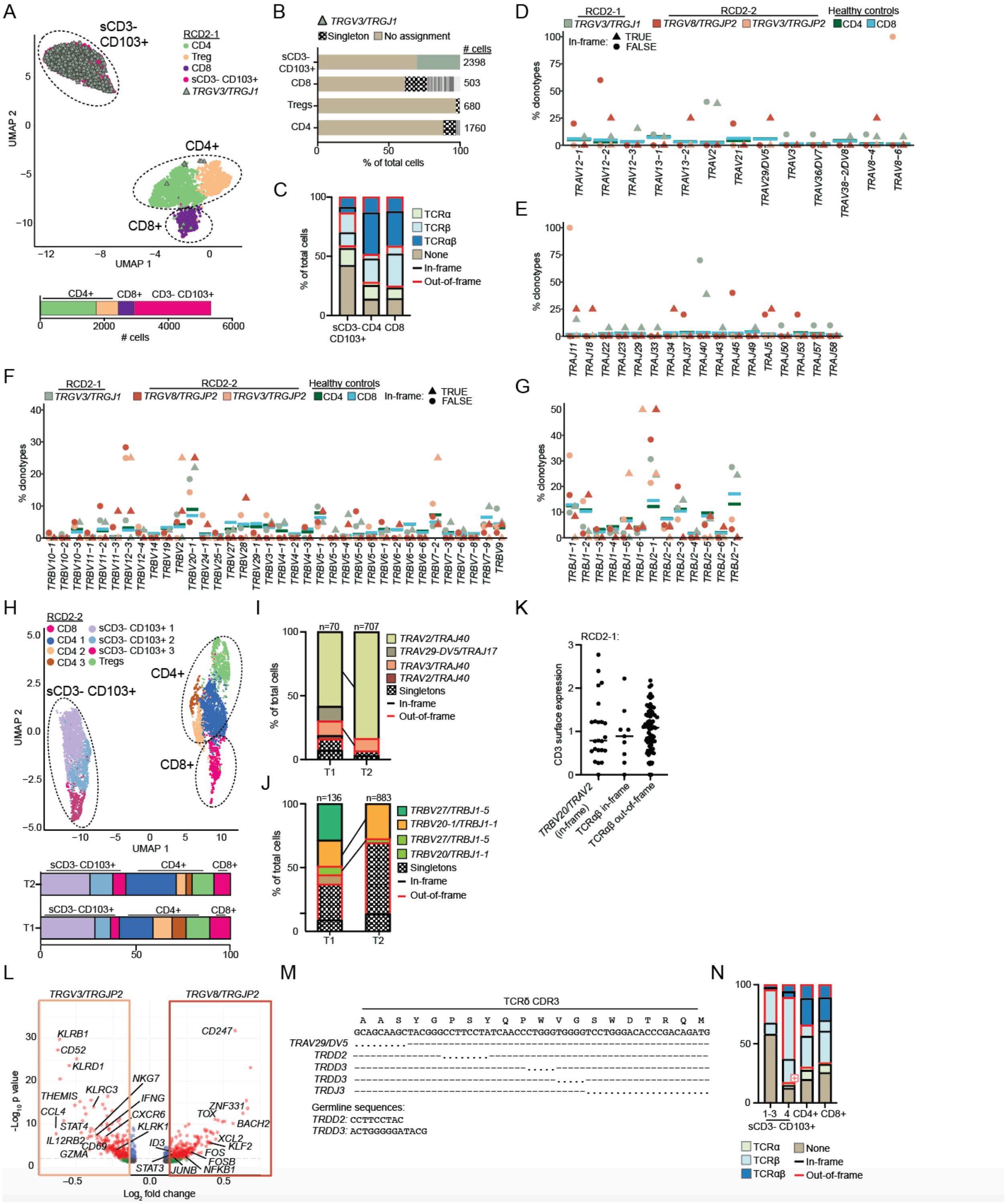


**Fig. S4. TCR analysis of RCD2 sCD3- CD103+ clones**

(**A**) UMAP analysis of CD45+ sCD3+ and sCD3- CD103+ cells for individual RCD2-1 (n=4,661 total cells) using combined gene-expression and cell surface protein data. The proportion of cell types are shown below the UMAP. Cells assigned the clonal TCRγ chain sequence

*TRGV3*/*TRGJP1* (n=730 cells) are highlighted on the UMAP. (**B**) Assignment of unique TCRγ V(D)J clonotypes to the cell types identified in the UMAP for individual RCD2-1. The *TRGV3*/*TRGJP1* clonal sequence is highlighted. Singletons refer to one cell carrying a unique TCRγ clonotype. Grey colors indicate TCRγ clones >1 cell. n=1160 total number of cells assigned a TCRγ chain. (**C**) Proportion of sCD3- CD103+, CD4+ or CD8+ cells that are assigned TCRα, TCRβ, or paired TCRα and TCRβ chains that are either in-frame or out-of-frame for individual RCD2-1. Number of cells: sCD3- CD103+, 2,398; CD4+, 1,760; CD8+, 530. (**D-G**) Variable (V) and Joining (J) gene usage of TCRα clonotypes (*TRAV*, D; *TRAJ*, E) and TCRβ clonotypes (*TRBV*, F; *TRBJ*, G) expressing unique V(D)J sequences from sCD3- CD103+ clones carrying the indicated clonal TCRγ sequence for individuals RCD2-1 and RCD2-2, and CD4 and CD8 T cells from healthy donors. TCRα and TCRβ clonotypes expressed by TCRγ sCD3- CD103+ clones are annotated as in-frame (triangles) or out-of-frame (circles), while clonotypes from healthy donor CD4 and CD8 cells are denoted as horizontal lines. (**H**) UMAP analysis of CD45+ sCD3+ and sCD3- CD103+ cells of two integrated timepoints ("T1" and "T2") from individual RCD2-1 (RCD2-1a and RCD2-1b) using combined gene-expression and cell surface protein data (n=6,110 total cells; T1, 575 cells; T2, 5,535 cells). T2 (RCD2-1b) is the initial sample presented, while T1 (RCD2-1a) is a biopsy taken four months prior. The proportion of cell types are shown below the UMAP. (**I-J**) Proportion of sCD3+ CD103- cells assigned the clonal *TRGV3*/*TRGJP1* TCRγ chain carrying unique TCRα (**I**) or TCRβ (**J**) V(D)J clonotypes. Expanded clonotypes (>1 cell) are highlighted as colors while singletons refer to one cell carrying a unique TCR sequence. (**K**) Expression (normalized reads) of surface CD3 protein of sCD3- CD103+ cells expressing the clonal TCRγ chain *TRGV3*/*TRGJP1* amongst cells assigned paired TCRα and TCRβ chains that are either in-frame or out-of-frame for individual RCD2-1. The dominant *TRBV20/TRAV2* in-frame clonotype is highlighted (n=23 cells), along with the remaining in-frame (n=9 cells) and out-of-frame (n=70 cells) TCRαβ clonotypes. (**L**) Volcano plot of differentially expressed genes between sCD3- CD103+ cells expressing *TRGV3*/*TRGJP2* (n=42 cells) and sCD3- CD103+ cells expressing *TRGV8*/*TRGJP2* (n=182 cells) for individual RCD2-2. Vertical dotted lines represent $\log_2$ fold change > 0.1. Genes that are significant are denoted in red and have a $\log_2$ fold change > 0.1 and p-value < 0.05. The full list of differentially expressed proteins and genes is described in data S3. (**M**) Amino acid sequence alignment of the TCRδ CDR3 region of the *TRAV29/DV5/TRDJ3* clone expressed by sCD3- CD103+ cells in individual RCD2-2. Alignment is shown against germline *TRAV29/DV5*, *TRDD* and *TRDJ3* genes. Dots indicate a sequence match, while dashed lines indicate no match. (**N**) Proportion of sCD3- CD103+, CD4+ or CD8+ cells that are assigned TCRα, TCRβ, or paired TCRα and TCRβ chains that are either in-frame or out-of-frame for individual RCD2-2. sCD3- CD103+ 1-3 clusters correspond to cells carrying the *TRGV3*/*TRGJP2* TCRγ chain while sCD3- CD103+ 4 cluster corresponds to cells carrying the *TRGV8*/*TRGJP2* TCRγ chain. Number of cells: sCD3- CD103+ 1-3, 421; sCD3- CD103+, 83; CD4+, 2,209; CD8+, 676.

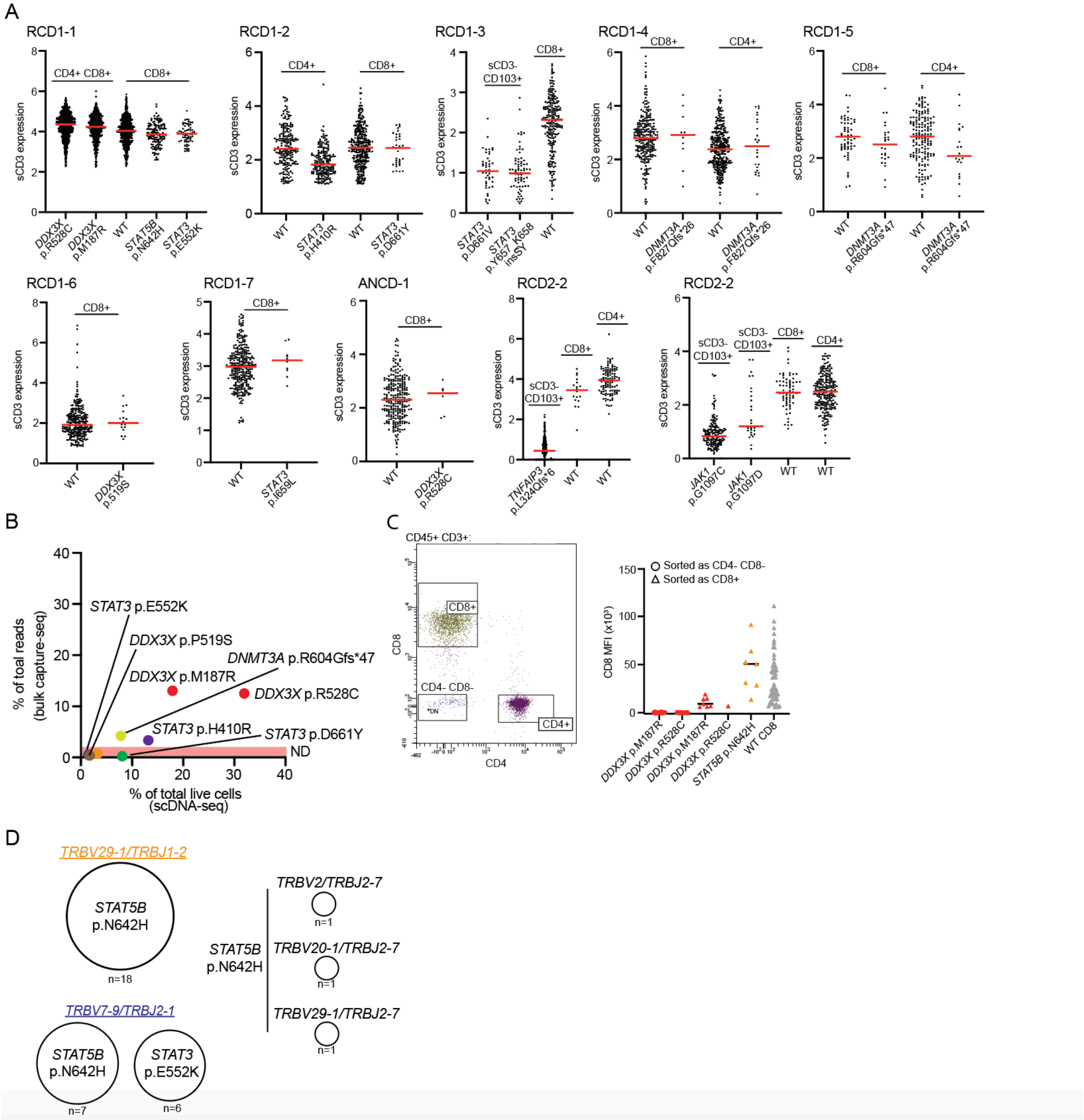


**Fig. S5. Surface CD3 expression and TCR analysis of sCD3+ T cell clones carrying mutations** (**A**) Expression (normalized reads) of surface CD3 from scDNA-Seq cell surface protein data of cells carrying the indicated mutation and cells that are wild-type (WT) for the indicated sample. Red lines indicate the median. The cell type is denoted above the WT and mutant cells. (**B**) Comparison of variants identified in scDNA-Seq with targeted bulk capture sequencing of the same biopsy for individuals RCD1-1 (*DDX3X* p.M187R, *DDX3X* p.R528C, *STAT3* p.E552K and *STAT5B* p.N642H), RCD1-2 (*STAT3* p.H410R and *STAT3* p.D661Y), RCD1-5 (DNMT3A p.R604Gfs*47) and RCD1-6 (*DDX3X* p.P519S). The highlighted red area indicates the approximate cut-off of the variant caller for calling variants from the targeted bulk sequencing. ND, not detected. (**C**) Flow cytometry index analysis of CD8 fluorescence intensity (right panel)

of CD45+ CD3+ cells sorted as either CD4- CD8- (circles) or CD8+ (triangles) with the indicated somatic mutation determined by G&T-Seq for individual RCD1-1. Left panel depicts the flow cytometry sorting strategy. n=110 total cells (26 *DDX3X* p.M187R CD4- CD8- cells, 17 *DDX3X* p.R528C CD4- CD8- cells, 7 *DDX3X* p.M187R CD8+ cells, 1 *DDX3X* p.R528C CD8+ cell, 7 *STAT5B* p.N642H CD8+ cells, 52 WT CD8+ cells. (**D**) Number of CD45+ CD3+ CD8+ cells sequenced by G&T-Seq for individual RCD1-1 that have either *STAT5B* p.N642H or *STAT3* p.E552K mutation calls and the indicated TCRαβ V(D)J sequences assigned.

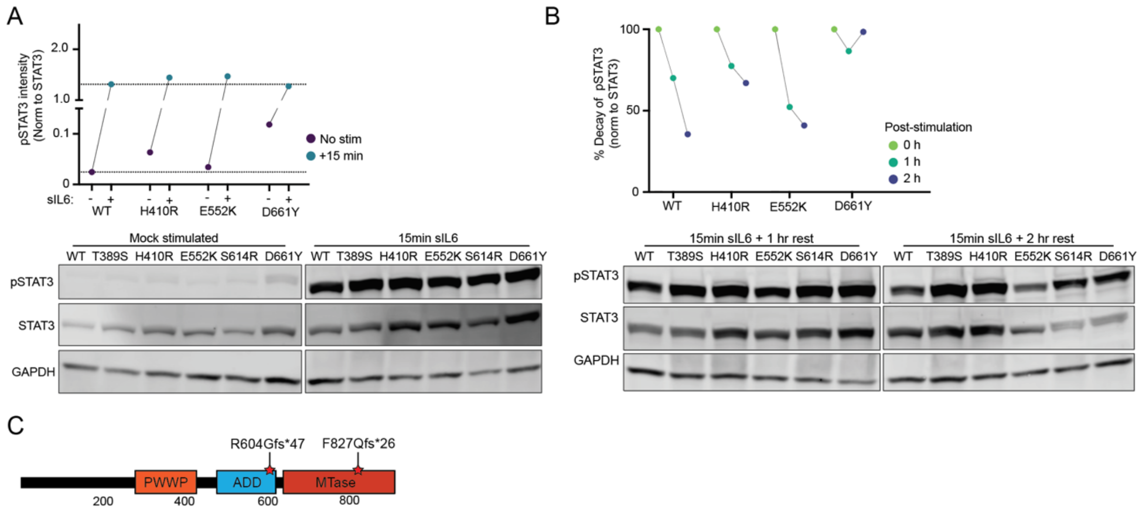


**Fig. S6. Functional characterization of *STAT3* mutations carried by sCD3+ T cell clones**
(**A-B**) Immunoblot of A4 STAT3$^{-/-}$ cells transfected with wild-type (WT) *STAT3* or the indicated *STAT3* variant with and without (mock) stimulation with sIL-6 for 15 min (A), or 1h or 2h following removal of stimulation (B). Top plots indicate the quantification of protein expression of the immunoblot depicted below. T389S and S614R are known germline and somatic gain-of-function mutations. Data was normalized to WT STAT3. GAPDH was used as a loading control. Experiment was repeated twice. (**C**) DNMT3A protein domains and location of somatic mutations identified in this study. PWWP, Pro-Trp-Trp-Pro. MTase, methyltransferase.

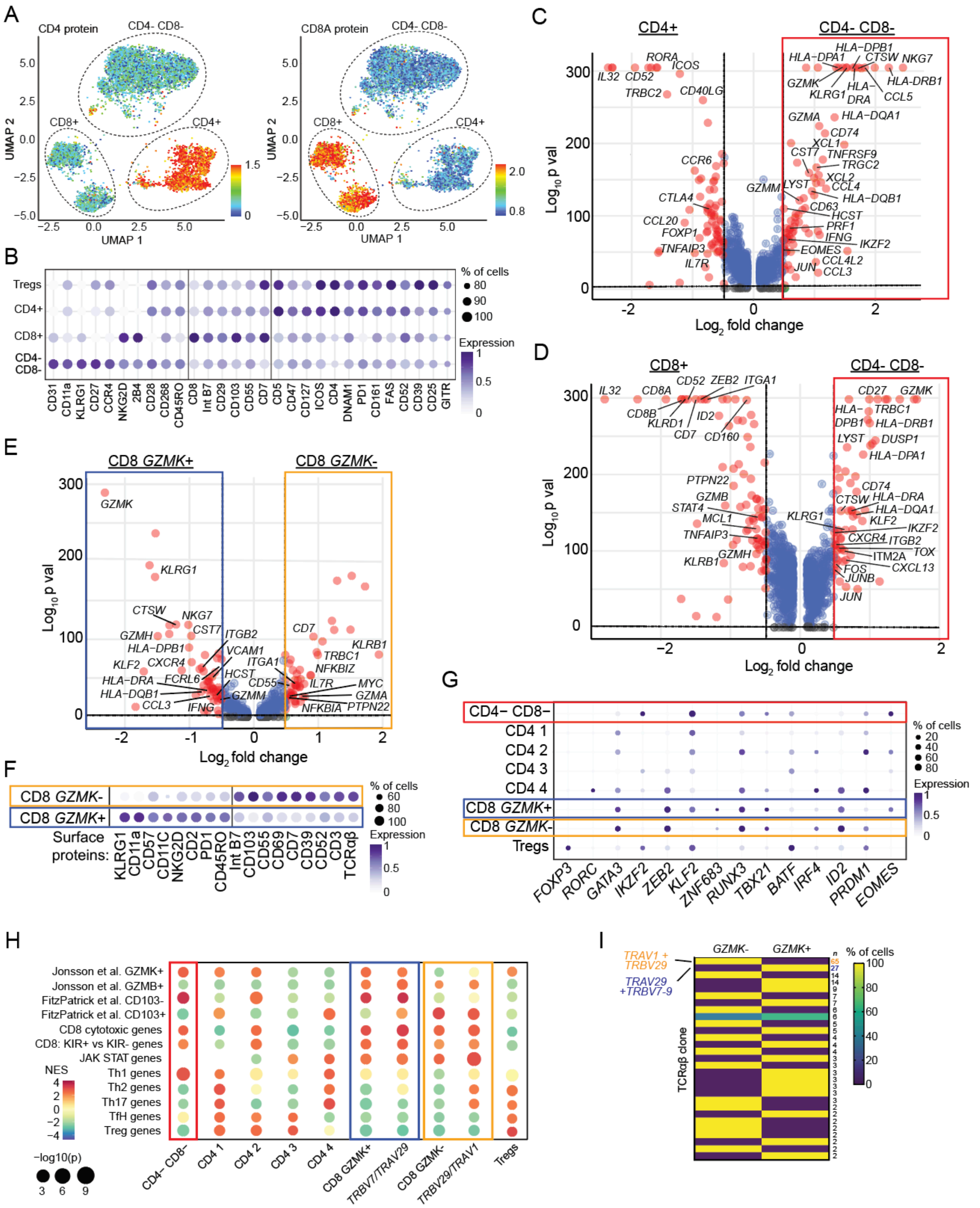


**Fig. S7. scRNA-Seq analysis of mutated T cell clones in individual RCD1-1**

(**A**) UMAP of CD45+ sCD3+ cells from individual RCD1-1 (n=7,569 cells, Fig. 5A) overlayed with protein expression of CD4 (left panel) and CD8 (right panel). (**B**) Dot plots of the top differentially expressed (p-value < 0.05) cell surface proteins between Tregs, CD4+, CD8+ and CD4- CD8- cell types. (**C-E**) Volcano plots of differentially expressed genes between: CD4+ (n=1,129) and CD4- CD8- (n=4,116) cells (C); CD8+ (n=1,707) and CD4- CD8- (n=4,116) cells (D); CD8+ GZMK+ (n=983) and CD8+ GZMK- (n=724) cells (E). Genes that are significant are denoted in red ($log_2$ fold change >0.1 and p-value <0.05), while those not significant are denoted in blue. Colored boxes mark cell types that contain TCRαβ T cell clones carrying somatic mutations detailed in Fig. 5. The full list of differentially expressed proteins and genes is described in data S3. (**F**) Dot plots of the top differentially expressed (p-value < 0.05) cell surface proteins between CD8+ GZMK- and CD8+ GZMK+ cells. (**G**) Dot plots of key T cell transcription factor genes amongst the different cell types indicated. (**H**) Gene set enrichment analysis (GSEA) utilizing a curated list of gene signatures (detailed in data S6) amongst the indicated cell type. The color intensity of each dot indicates the relative enrichment (NES) of that signature compared to other cell types. NES: Normalized Enrichment Score. (**I**) CD8 GZMK- and CD8 GZMK+ cell type annotation of expanded sCD3+ CD8+ TCRαβ clones (>1 cell, n=29 clones). The number of cells belonging to each clone is indicated along with the *STAT3* and *STAT5B* mutated CD8 T cell clones. For the dotplots depicted in (B) and (F to G) the size of each dot represents the percentage of cells with non-zero expression, and color intensity represents scaled log expression averaged over all cells.

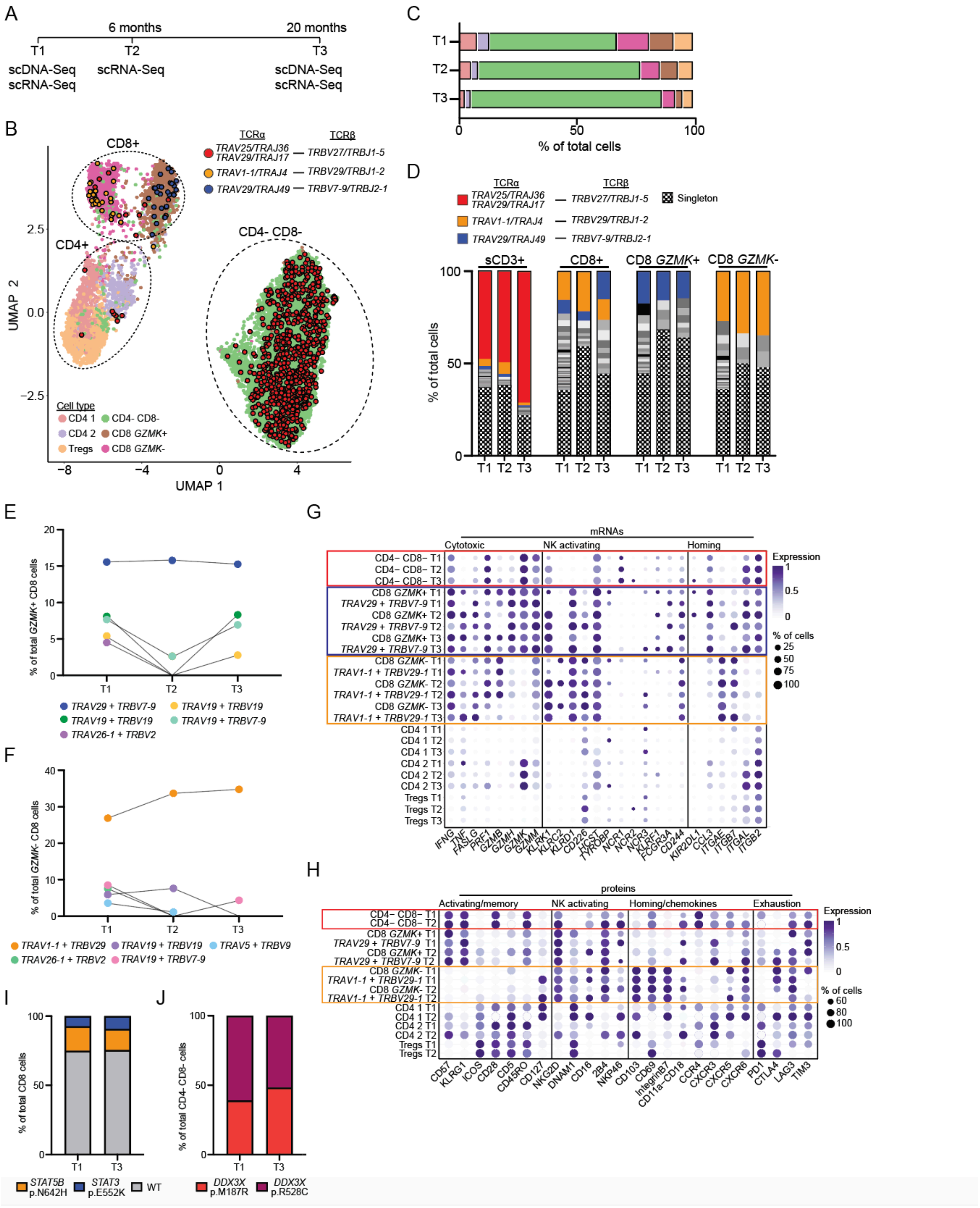


**Fig. S8. Longitudinal analysis of mutant TCR+ clones**

(**A**) Schematic of the longitudinal samples and single cell technology used for individual RCD1-1 (T1, RCD1-1a; T2, RCD1-1b; T3, RCD1-1c). T1 represents the original sample presented in this study. (**B**) UMAP of CD45+ sCD3+ cells from individual RCD1-1 (n=12,520 cells) using combined gene-expression and cell surface protein data of the three timepoints integrated together. Clusters are denoted by colors and labelled by cell type. The indicated TCRαβ chain clones are highlighted in the UMAP. (**C**) Proportion of cell types identified in the UMAP analysis amongst the different timepoints. Cell type colors correspond to those depicted in (B). (**D)** Proportion of cells assigned paired TCRα and TCRβ chains carrying unique V(D)J sequences amongst the indicated cell type and timepoint (n=1,832 total TCRαβ cells). TCRαβ clonal sequences corresponding to those with somatic mutations are shown. Singletons refer to one cell carrying a unique TCRαβ sequence. Grey colors indicate unique TCRαβ clones >1 cell. (**E-F**) Frequency of the top five expanded TCRαβ clones amongst CD8 GZMK+ cells (E) and CD8 GZMK- cells (F) for each timepoint. Number of CD8 GZMK+ cells: T1, 806 cells; T2, 253 cells; T3, 46 cells. Number of CD8 GZMK- cells: T1, 1063, T2, 277 cells; T3, 85 cells. (**G-H**) Dot plot of selected genes (G) and proteins (H) for the indicated cell type and time point. Colored boxes mark cells, or the cell type they belong to, of the TCRαβ sequences of mutated T cell clones. The size of each dot represents the percentage of cells with non-zero expression. The color intensity represents the scaled log expression averaged over all cells. (**I**) Frequency of CD8+ cells that are mutant for *STAT5B* p.N642H or *STAT3* p.E552K*,* or wild-type (WT) for the indicated timepoints from scDNA-Seq data. Number of CD8 cells: T1, 830 cells; T3, 507 cells. (**J**) Frequency of CD4- CD8- cells that are mutant for *DDX3X* p.M187R or *DDX3X* p.R528C cells for the indicated timepoints from scDNA-Seq data. Number of CD4- CD8- cells: T1, 2504 cells; T3, 1359 cells.

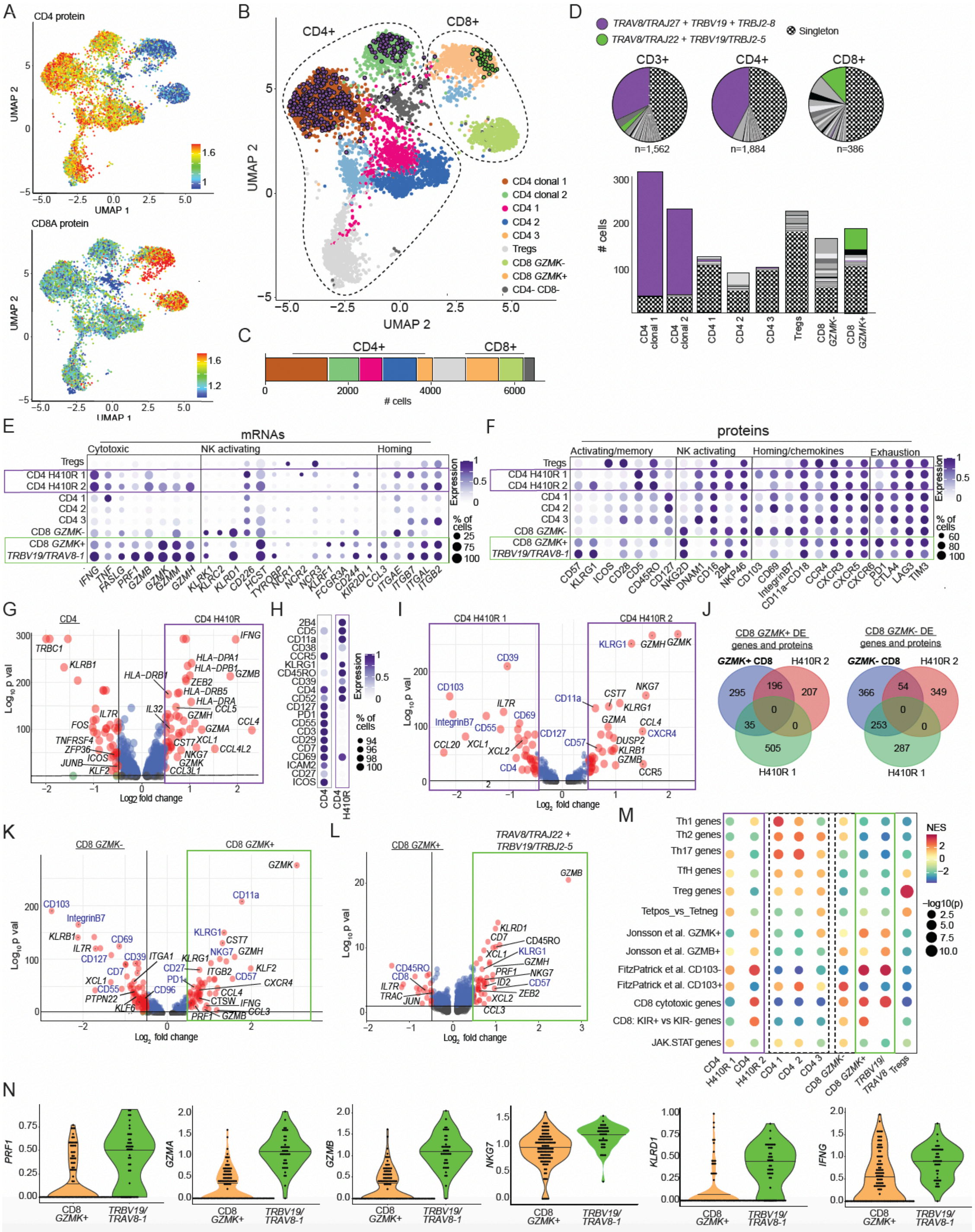


**Fig. S9. scRNA-Seq analysis of mutated T cell clones in individual RCD1-2**

(**A-B**) UMAPs of CD45+ sCD3+ cells from individual RCD1-2 (n=6,525 cells) using combined gene-expression and cell surface protein data. Panel A shows protein expression of CD4 (top) and CD8 (bottom). Panel B shows the cell types assigned to different clusters and TCRαβ clones described in (D). (**C**) The proportion of cell types identified from the UMAP analysis. (**D**) Proportion of cells assigned paired TCRα and TCRβ chains carrying unique V(D)J sequences amongst the indicated cell type (n=3,832 total TCRαβ assigned cells). TCRαβ clonal sequences corresponding to those with somatic driver mutations are shown. Singletons refer to one cell carrying a unique TCRαβ sequence. Grey colors indicate unique TCRαβ clones >1 cell. (**E-F**) Dot plot of selected genes (E) and proteins (F) for the indicated cell type. Colored boxes mark cells, or the cell type they belong to, of selected TCRαβ sequences of mutated T cell clones. The size of each dot represents the percentage of cells with non-zero expression. The color intensity represents the scaled log expression averaged over all cells. (**G**) Volcano plot of differentially expressed genes between nonclonal CD4 cells (clusters CD4 1, CD4 2 and CD4 3, n=1,773) and CD4 *STAT3* H410R clonal cells (clusters CD4 H410R 1 and CD4 H410R 2, n=2,291). (**H**) Dot plots of cell surface proteins that are significantly differentially expressed (p-value < 0.05) between CD4 cells and CD4 H410R cells. (**I**) Volcano plot of differentially expressed genes and proteins between CD4 H410R 1 (n=1,537) and CD4 H410R 2 (n=754) clusters. (**J**) Overlap of genes and proteins that are differentially expressed in either CD8 GZMK+ (left panel) or CD8 GZMK- cells (right panel) with genes and protein differentially expressed between CD4 H410R 1 and CD4 H410R 2 clusters. (**K-L**) Volcano plots of differentially expressed genes and proteins between: CD8+ GZMK- (n=590) and CD8+ GZMK+ (n=798) clusters (K); CD8 GZMK+ cells assigned paired TCRαβ polyclonal V(D)J sequences (n=146) and CD8 GZMK+ cells assigned paired TCRαβ V(D)J sequences containing *TRAV8/TRAJ22* and *TRBV19/TRBJ2-5* (n=44) (L). (**M**) GSEA of curated gene sets (detailed in data S6) amongst the indicated cell type. The color intensity of each dot indicates the relative enrichment (NES) of that signature compared to other cell types. NES: Normalized Enrichment Score. (**N**) Violin plots of the expression (normalized counts) of selected genes of CD8 GZMK+ cells assigned TCRαβ polyclonal V(D)J sequences or CD8 GZMK+ cells assigned *TRAV8/TRAJ22* and *TRBV19/TRBJ2-5*. Horizontal lines in each violin represents the mean. The full list of differentially expressed proteins and genes for the volcano plots are described in data S3.

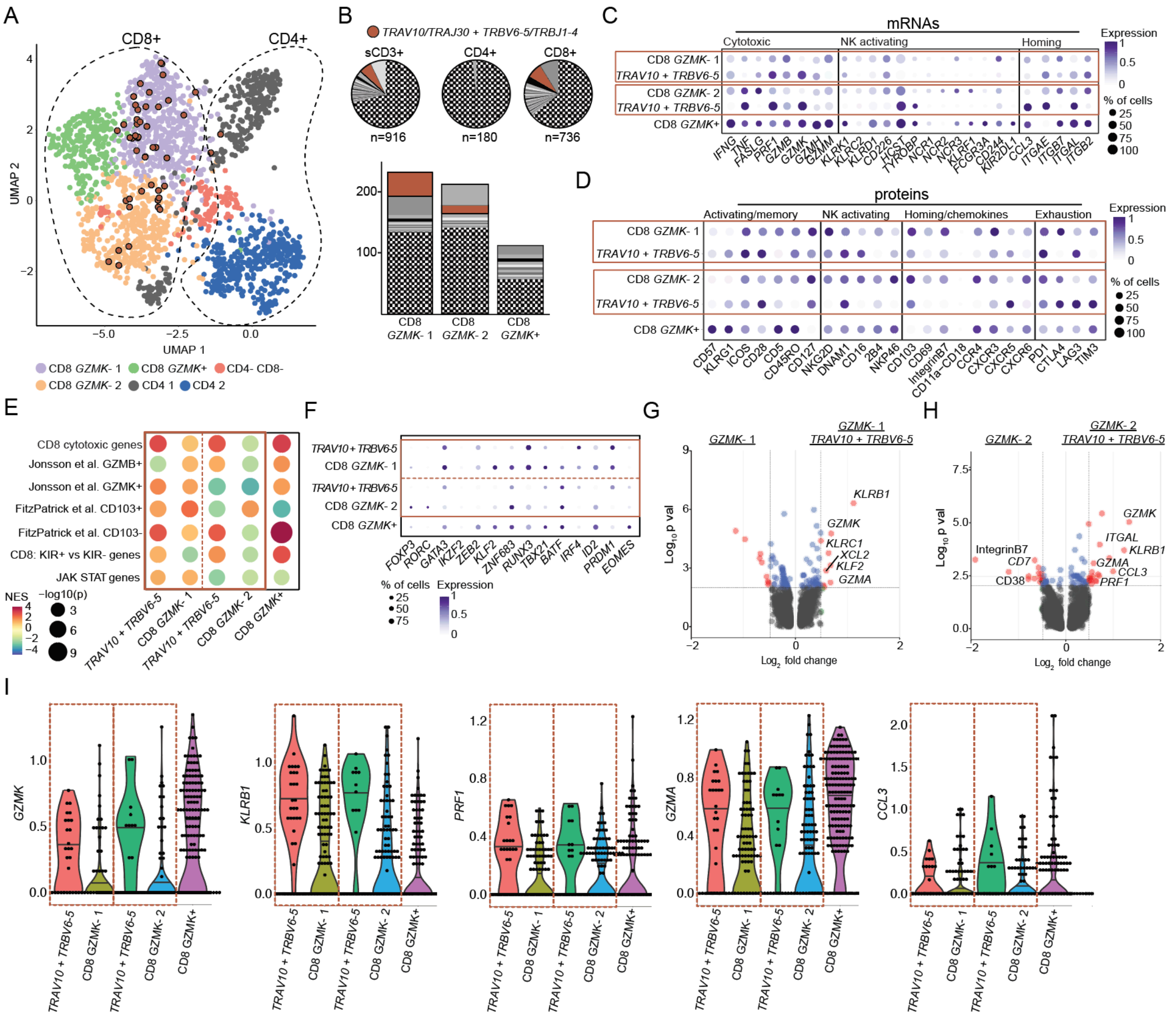


**Fig. S10. scRNA-Seq analysis of mutated T cell clones in individual RCD1-6**

(A) UMAP of CD45+ sCD3+ cells from individual RCD1-6 (n=1,963 cells) using combined gene-expression and cell surface protein data. Clusters are denoted by colors and labelled by cell type. The TCRαβ clone denoted in (B) is highlighted in the UMAP. (**B**) Proportion of cells assigned TCRα and TCRβ chains carrying unique V(D)J sequences amongst the indicated cell type (n=1,832 total TCRαβ cells). TCRαβ clonal sequences corresponding to those with somatic driver mutations are shown (*DDX3X* P519S). Singletons refer to one cell carrying a unique TCRαβ sequence. Grey colors indicate unique TCRαβ clones >1 cell. (**C-D**) Dot plots of selected genes (C) and proteins (D) for the indicated cell type. Colored boxes mark cells, or the cell type they belong to, of the TCRαβ sequences of *DDX3X* P519S mutated T cell clone. The size of each dot represents the percentage of cells with non-zero expression. The color intensity represents the scaled log expression averaged over all cells. (**E**) GSEA of curated gene sets (detailed in data S6) amongst the indicated cell type. The color intensity of each dot indicates the relative enrichment (NES) of that signature compared to other cell types. NES, Normalized Enrichment Score. (**F**) Dot plot of the expression of selected transcription factor genes amongst the indicated cell types. (**G-**

**H**) Volcano plots of differentially expressed genes and proteins between: CD8+ *GZMK*- 1 cells assigned TCRαβ polyclonal V(D)J sequences (n=300 cells) and CD8 *GZMK*- 1 cells assigned TCRαβ V(D)J sequences containing *TRAV10/TRAJ30* and *TRBV6-5/TRBJ1-4* (n=15 cells) (G), CD8+ *GZMK*- 2 cells assigned TCRαβ polyclonal V(D)J sequences (n=237 cells) and CD8 *GZMK*- 2 cells assigned TCRαβ V(D)J sequences containing *TRAV10/TRAJ30* and *TRBV6-5/TRBJ1-4* (n=15 cells) (H). The full list of differentially expressed proteins and genes for the volcano plots is described in data S3. (**I**) Violin plots of the expression (normalized counts) of selected genes of the indicated CD8 cell types assigned paired TCRαβ V(D)J sequences, separated for cells expressing *TRAV10/TRAJ30* and *TRBV6-5/TRBJ1-4* (marking *DDX3X* P519S mutant cells). Horizontal lines in each violin represents the mean.

### Table S1. Patient metadata

| Disease type | Sample | Initial presenting symtpoms | Duration of symptoms prior to CD dianosis | RCD symptoms | Years since diagnosis of CD | Sex | Comorbidities | HLA-DQ (and zygosity when known) | Duodenal histology (Marsh score) | Immunohistochemistry | Flow cytometry | Celiac serology at time of biopsy |
|---|---|---|---|---|---|---|---|---|---|---|---|---|
| Healthy control | C-1 | Asymptomatic Abnormal imaging | NA | NA | NA | M | Nil remarkable | Not performed | 0 | Not performed | Not performed | Not performed |
| | C-2 | Gastro-oesophageal reflux disease (GORD) | NA | NA | NA | M | Hypertension, obstructive sleep apnea | Not performed | 0 | Not performed | Not performed | TTG <1, DGP <1 |
| | C-3 | GORD | NA | NA | NA | M | Hypercholesterolaemia | Not performed | 0 | Not performed | Not performed | Not performed |
| | C-4 | GORD | NA | NA | NA | M | Asthma | Not performed | 0 | Not performed | Not performed | Not performed |
| | C-5 | GI bleeding | NA | NA | NA | M | Nil remarkable | Not performed | 0 | Not performed | Not performed | Not performed |
| | C-6 | Dysphagia | NA | NA | NA | F | Asthma | Not performed | 0 | Not performed | Not performed | Not performed |
| | C-7 (treated CD) | Bloating, fatigue | < 5 years | NA | 2 | M | Lichen planopilaris | DQ2.5 | 0 | Not performed | Not performed | NEG |
| Active, newly diagnosed celiac disease | ANCD-1 | Iron deficiency, bloating | < 6 months | NA | 0 | M | Type 1 diabetes | Not performed | 3A | Not performed | Not performed | TTG >250 (<15) |
| | ANCD-2 | Iron deficiency, abdominal pain | < 5 years | NA | 0 | F | Nil remarkable | DQ2.5 Homozygous | 3C | Not performed | Not performed | TTG 98 (<20), DGP 60 (<20) |
| | ANCD-3 | Iron deficiency, lethargy constipation | 7 years | NA | 0 | F | Nil remarkable | DQ2.5 Heterozygous | 3B | Not performed | Not performed | TTG 6 (<4), DGP 36 (<20) |
| | ANCD-4 | Bloating, abdominal pain | 12 months | NA | 0 | F | Nil remarkable | DQ2.5 Homozygous | 3C | Not performed | Not performed | TTG 197 (<20), DGP 49 (<20) |
| RCD type 1 | RCD1-1a | Difficulty gaining weight, mild bloating | < 5 years | Low grade GI upset | 12 | M | Nil remarkable | DQ2.5 | 3A | Normal; most IELs are CD8+ | Abnormal, uncertain significance: atypical infiltrate of small lymphocytes in the lamina propria with 54% being CD2+, sCD3+, CD4-, CD5+, CD7+/-, CD8-, CD10- and CD16/56-. No aberrant IEL sCD3- CD103+ clones detected. | NEG |
| | RCD1-1b | | | | 12 | | Nil remarkable | | 3B | Normal; most IELs are CD8+ | 82% lymphocytes are T cells (CD4/CD8 ratio 0.6); 39% of T cells are CD3+ CD4- and CD8-. No aberrant IEL sCD3- CD103+ clones detected. | NEG |
| | RCD1-1c | | | | 14 | | Nil remarkable | | 3C | CD2 positive, with CD4+ > CD8+ T cells | No aberrant IEL sCD3- CD103+ clones detected. | NEG |
| | RCD1-2 | Diarrhea, 20 kg weight loss | < 5 years | Ongoing poor weight and lethargy from diagnosis | 2 | M | Osteoporosis, pancreatic insufficiency, abnormal LFTs, immune thrombocytopenic purpura | DQ2.2/DQ8 | 3C | Normal | Abnormal but significance uncertain: 39% of the CD3+ T cells are CD2+, CD3+, CD5+ (dim), CD7+, CD16/56-, TCD gd- and CD4/CD8 ratio 0.8 | NEG |
| | RCD1-3 | Diarrhea, 8 kg weight loss, abdominal pain, anorexia | ~30 years | Ongoing abdominal pain from diagnosis | 1 | M | Hypertension | DQ2.5 | 3B | Normal | Normal | Low POS (TTG 26, n<20) |
| | RCD1-4 | Diarrhea, 11kg weight loss, abdominal pain, lethargy | < 5 years | Ongoing abdominal pain and diarrhea from diagnosis | 1 | M | Osteopaenia, neutropaenia, central hypogonadism | DQ2.5 Homozygous | 3A | Normal | Normal | NEG |
| | RCD1-5 | Screening | NA | Iron deficiency, GI upset | 38 | F | Psoriatic arthritis, primary billary cirrhosis, osteoporosis | DQ2.5/DQ8 | 3C | Normal | Normal | NEG |
| | RCD1-6 | Iron deficiency, GI Upset | < 5 years | Lethargy, brain fog, GI upset | 4 | F | Osteopaenia | DQ2.5 Heterozygous | 3C | Normal | Normal | NEG |
| | RCD1-7 | Diarrhea | < 5 years | Diarrhea, fatigue, low iron | 18 | F | Rosacea, asthma | Not performed | 1 (Following 3 months topical steroid therapy) | Normal | Normal | NEG |
| | RCD1-8 | GI Upset | < 5 years | Diarrhea, weight loss | 9 | F | Lymphocytic colitis | Not performed | 3B | Normal | Not performed | NEG |
| | RCD1-9 | Iron deficiency | < 5 years | Diarrhea | 14 | F | Microscopic colitis | DQ2.5 | 3C (primarily in bulb) | Normal | Normal | NEG |
| | RCCD1-10 | Diarrhoea, 12 kg weight loss, abdominal pain | < 5 years | Abdominal cramps, urgency, diarrhea, low iron | 26 | M | Osteopaenia | Not performed | 1 (Following 3 months topical steroid therapy) | Normal | Sample insufficient | NEG |
| RCD type 2 | RCD2-1a | Diarrhoea | < 5 years | Diarrhoea, 13kg weight loss | 14 | M | Hypertension, gout | DQ2.5 | 3C | Abnormal: CD3+ IEL have >50% loss of CD8 expression | Abnormal: >20% sCD3- CD103+ CD8- IELs | NEG |
| | RCD2-1b | | | | 14 | | Hypertension, gout | | 3C (Following SC Cladribine) | Unchanged | Abnormal clone still present | NEG |
| | RCD2-2 | Abdominal pain, diarrhoea, weight loss (>10kg) | 3 months | Diarrhoea, weight loss, hypoalbuminaemia, peripheral oedema, electrolyte imbalance, iron deficiency anaemia | 0 | M | None (multiple nutritional deficiences as a consequence of RCD2) | DQ2.5 | 3B | Abnormal: IEL positivity for CD3 and CD7, negativity for CD4, CD5, CD8, no expression of CD30 and CD56 | Abnormal: >20% sCD3- cyCD3+ CD103+ CD7+ IELs | TTG IgA 103 (<20) EMA positive 1/40 |

Table S2. Variants identified from scDNA-Seq

| Variant | Genome coordinate (Hg38) | Sample | Polyphen score | Sift score | CADD score | B cells | CD4+ cells | CD8+ cells | Epithelial cells | CD4- CD8- cells | sCD3+ CD103+ cells |
|---|---|---|---|---|---|---|---|---|---|---|---|
| *DDX3X* p.Thr532Ala | chrX:41346601 C>T | RCD2-1 | 0 | 0 | 25.2 | 16/282 | 4/104 | 3/17 | 6/212 | NA | 308/309 |
| *TNFAIP3* p.Leu324Glnfs*6 | chr6:137877235-137877239 -CTCAT | RCD2-1 | NA | NA | NA | 14/282 | 3/104 | 3/17 | 5/212 | NA | 303/309 |
| *KMT2D* p.Gln2380* | chr12:49040632 G>A | RCD2-1 | NA | NA | 45 | 11/282 | 3/104 | 1/17 | 1/212 | NA | 286/309 |
| *POT1* p.Gly236Val | chr7:124853134 C>A | RCD2-1 | 0 | 0 | 25 | 12/282 | 1/104 | 1/17 | 3/212 | NA | 232/309 |
| *JAK1* p.Gly1097Cys | chr1:64835476 C>A | RCD2-2 | 0 | 0 | 26.9 | 2/104 | 3/203 | 2/60 | 0/131 | NA | 147/225 |
| *KMT2D* p.Cys1408Phe | chr12:49047978 C>A | RCD2-2 | 0 | 0 | 27.5 | 2/104 | 3/203 | 2/60 | 1/131 | NA | 134/225 |
| *TET2* p.Asn1118Lysfs*11 | chr4:105237289 +A | RCD2-2 | NA | NA | NA | 3/104 | 2/203 | 2/60 | 0/131 | NA | 137/225 |
| *JAK1* p.Gly1097Asp | chr1:64835476 C>T | RCD2-2 | 0 | 0 | 27.7 | 0/104 | 1/203 | 1/60 | 0/131 | NA | 15/225 |
| *JAK3* p.Met511Ile | chr19:17838299 C>T | RCD2-2 | 0.17 | 0.17 | 22.6 | 0/104 | 2/203 | 1/60 | 0/131 | NA | 13/225 |
| *DDX3X* p.Arg528Cys | chrX:41346589 C>T | RCD1-1 | 0.02 | 0.02 | 28.2 | 7/420 | 55/752 | 86/832 | NA | 1305/2511 | NA |
| *DDX3X* p.Met187Arg | chrX:41343232 T>G | RCD1-1 | 0 | 0 | 26.7 | 3/420 | 26/752 | 59/832 | NA | 727/2511 | NA |
| *SOCS3* p.Val84Ile | chr17:78358846 C>T | RCD1-1 | 0.06 | 0.06 | 23.4 | 0/420 | 21/752 | 52/832 | NA | 512/2511 | NA |
| *STAT5B* p.Asn642His | chr17:42207711 T>G | RCD1-1 | 0.09 | 0.09 | 25.5 | 0/420 | 2/752 | 147/832 | NA | 1/2511 | NA |
| *STAT3* p.Glu552Lys | chr17:42323354 C>T | RCD1-1 | 0.01 | 0.01 | 25.4 | 0/420 | 1/752 | 66/832 | NA | 1/2511 | NA |
| *STAT3* p.His410Arg | chr17:42329558 T>C | RCD1-2 | 0 | 0 | 26.9 | 4/299 | 232/538 | 4/470 | 0/279 | NA | NA |
| *STAT3* p.Asp661Tyr | chr17:42322402 C>A | RCD1-2 | 0.03 | 0.03 | 26.7 | 0/299 | 1/538 | 37/470 | 0/279 | NA | NA |
| *STAT3* p.Asp661Val | chr17:42322401 T>A | RCD1-3 | 0.43 | 0.43 | 26.8 | 0/144 | NA | 4/227 | 4/484 | NA | 41/147 |
| *TET2* p.His1036Leufs*9 | chr4:105237048-105237075 -CACGCCAAGTCGTTATTTGACCATAAGG | RCD1-3 | NA | NA | NA | 0/144 | NA | 3/227 | 11/484 | NA | 65/147 |
| *STAT3* p.Tyr657_Lys658insSerTyr | chr17:42322410 +TATAGC | RCD1-3 | NA | NA | NA | 1/144 | NA | 7/227 | 10/484 | NA | 67/147 |
| *DNMT3A* p.Phe827Glnfs*26 | chr2:25235821-25235825 -CTGAA | RCD1-4 | NA | NA | NA | 21/155 | 24/333 | 10/275 | 2/207 | NA | NA |
| *DNMT3A* p.Arg604Glyfs*47 | chr2:25244196 -G | RCD1-5 | NA | NA | NA | 10/174 | 20/194 | 26/88 | 8/569 | 29/118 | NA |
| *DDX3X* p.Pro519Ser | chrX:41346562 C>T | RCD1-6 | 0 | 0 | 25.6 | 0/209 | 0/104 | 19/318 | 0/418 | NA | NA |
| *STAT3* p.Ile659Leu | chr17:42322408 T>G | RCD1-7 | 0.25 | 0.25 | 25.1 | 0/210 | 0/429 | 10/310 | 0/376 | NA | NA |
| *DDX3X* p.Arg528Cys | chrX:41346589 C>T | ANCD-1 | 0.02 | 0.02 | 28.2 | 0/343 | 0/362 | 6/245 | 0/375 | NA | NA |

#### Table S3. TCR sequences of mutated T cell clones

| Sample | Cell type | Mutations | *TRAV* | CDR3α (nucleotide) | CDR3α (protein) | *TRAJ* | *TRBV* | CDR3β (nucleotide) | CDR3β (protein) | *TRBJ* |
|---|---|---|---|---|---|---|---|---|---|---|
| RCD1-1 | CD4- CD8- | *DDX3X* M187R/ *DDX3X* R528C | *TRAV25*<br>*TRAV29/ DV5* | TGTGCAGGGCCGGTGAGTGGGGCAAACAACCTCTTCTTT<br>TGTGCAGCAAGCGCGATTCAAGCTGCAGGCAACAAGCTAACTTTT | CAGPVSGANNLF<br>CAASAIQAAGNKLTL | *TRAJ36*<br>*TRAJ17* | *TRBV27* | TGTGCCAGCAGTTATGTCAGGGGGACCCGGCCCCAGCATTTT | CASSYVRGTRPQHF | *TRBJ1-5* |
| RCD1-1 | CD8+ | *STAT5B* N642H | *TRAV1-1* | TGCGCTGTGATATCTGGTGGCTACAATAAGCTGATTTTT | CAVISGGYNKLIF | *TRAJ4* | *TRBV29-1* | TGCAGCGTTGAATGGGGGGACAGGGAAGGCTACACCTTC | CSVEWGDREGYTF | *TRBJ1-2* |
| RCD1-1 | CD8+ | *STAT3* E552K/ *STAT5B* N642H | *TRAV29/ DV5* | TGTGCAGCAGTGGCCACCGGTAACCAGTTCTATTTT | CAAVATGNQFYF | *TRAJ49* | *TRBV7-9* | TGTGCCAGCAGCCCTCAATATAAGCGACTAGCGGGAGGACCCGAGCAGTTCTTC | CASSPQYKRLAGGPEQFF | *TRBJ2-1* |
| RCD1-2 | CD4+ | *STAT3* H410R | *TRAV8-1* | TGTGCCGTGAAATCCGCTGGAGGGACCAATGCAGGCAAATCAACCTTT | CAVKSAGGTNAGKSTF | *TRAJ27* | *TRBV19* | TGTGCCAGTAGTTTTGAGGTCCAATCCTACGAGCAGTACTTC | CASSFEVQSYEQYF | *TRBJ2-7* |
| RCD1-2 | CD8+ | *STAT3* D661Y | *TRAV8-1* | TGTGCCGTGACCGCTTTTTCTGGTTCTGCAAGGCAACTGACCTTT | CAVTAFSGSARQLTF | *TRAJ22* | *TRBV19* | TGTGCCAGTAGTCAAGGACTAGCGGGACAAGAGACCCAGTACTTC | CASSQGLAGQETQYF | *TRBJ2-5* |
| RCD1-6 | CD8+ | *DDX3X* P519S | *TRAV10*<br>*TRAV26-1* | TGCGTGGTGACCATCACTCCCAGCTGCCGGCAGCATGCCCATTTT<br>TGCATCGTATTCAAAGCTGCAGGCAACAAGCTAACTTTT | CVVTITPSCRQHAHL<br>CIVFKAAGNKLTF | *TRAJ39*<br>*TRAJ17* | *TRBV6-5* | TGTGCCAGCAGAGCTTTTGACTCAACTAATGAAAAACTGTTTTTT | CASRAFDSTNEKLFF | *TRBJ1-4* |
